# An automated, explainable, NCCT-based clinical decision-support system for spontaneous intracerebral hemorrhage

**DOI:** 10.64898/2026.09.30.26364459

**Authors:** Chunyang Liu, Qi Shao, Yuchen Li, Chao Yuan, Huan Xiang, Yunhan Xu, Chenxi Yin, Jian Zhang, Yanchao Liang, Guang Yang

## Abstract

**Background:** Spontaneous intracerebral hemorrhage (ICH) has extremely high rates of disability and mortality. However, traditional CT imaging assessment relies solely on hematoma volume and rough anatomical location, making it difficult to account for prognostic heterogeneity and lacking precise individualized predictive tools. In recent years, radiomics has been able to extract texture and heterogeneity features from CT images at high throughput, and combined with machine learning algorithms, it has provided new possibilities for prognostic assessment.

**Methods:** In this retrospective, multicenter study, we included 2680 consecutive patients with spontaneous intracerebral hemorrhage, comprising a training cohort (n = 1876), an internal validation cohort (n = 804), and an independent external validation cohort (n = 196). We used deep learning algorithms to automatically segment hematomas and then extracted 1690 radiomics features from admission NCCT images. After reproducibility assessment, univariate screening, and LASSO feature selection, 42 valid radiomics features were integrated into a single radiomics feature spectrum (Rad-score). The Rad-score was combined with demographic features, clinical variables, and conventional CT radiomics features to construct two complementary machine learning models, used for functional outcome prediction and comprehensive prognostic stratification, respectively. Model performance was evaluated using R², mean squared error (MSE), area under the receiver operating characteristic (AUC), calibration analysis, decision curve analysis, and independent external validation. Furthermore, the possibility of ambient temperature variation at symptom onset as a prognostic modifier was explored.

**Results:** Among the 1,690 extracted radiomic features, 42 reproducible features were selected to construct the Rad-score. Incorporation of the radiomics signature substantially improved predictive performance compared with models based solely on conventional clinical and imaging variables. For functional outcome prediction, the integrated model increased the coefficient of determination (R²) from 0.35 to 0.93 while reducing the mean squared error (MSE) from 387 to 61. For prognostic risk stratification, the model based only on clinical variables achieved an AUC of 0.59, whereas the fully integrated model achieved an AUC of 0.95 and maintained robust performance in the external validation cohort (AUC=0.86).

**Conclusions:** We developed and validated a machine learning–based clinical decision support system integrating conventional CT radiomics and clinical variables for prognostic prediction and risk stratification in patients with ICH. By capturing quantitative imaging information beyond conventional CT markers, this framework significantly improved prognostic discrimination and demonstrated good generalizability across independent cohorts. These findings support the integration of quantitative imaging biomarkers into clinical decision-making for spontaneous ICH. Furthermore, the observed association between ambient temperature variation and disease progression warrants further investigation in prospective studies.

## Introduction

Spontaneous intracerebral hemorrhage (ICH) is one of the most devastating forms of stroke[1]. Although it accounts for only approximately 10–20% of all cerebrovascular events, it contributes disproportionately to stroke-related mortality and long-term disability[2, 3]. Despite substantial advances in neurocritical care and neurosurgical management, clinical outcomes remain highly heterogeneous[4, 5]. Patients with apparently similar hematoma volume, anatomical location, and neurological deficits often experience markedly different functional recovery and long-term prognosis[6]. Consequently, accurate early risk stratification remains one of the most important unmet clinical needs in the management of spontaneous ICH[6, 7].

Because of its widespread availability, rapid acquisition, and excellent sensitivity and specificity for acute hemorrhage, non-contrast computed tomography (NCCT) is the first-line imaging modality for the diagnosis and emergency evaluation of spontaneous ICH[8, 9]. Current clinical decision-making primarily relies on conventional CT findings and neurological severity scales. These variables form the basis of widely used prognostic scoring systems and surgical decision-making algorithms[10].However, these conventional indicators often fail to explain the significant differences in prognosis among patients with similar clinical presentations[11].

Recent advances in artificial intelligence have substantially improved the automatic detection and segmentation of intracranial hemorrhage on CT images[12, 13]. At the same time, radiomics, as a powerful quantitative imaging method, can extract hundreds to thousands of reproducible features. These high-dimensional imaging biomarkers demonstrate significant prognostic value in a variety of neurological and oncological diseases by capturing latent biological information that cannot be obtained through conventional visual assessment[14, 15]. Nevertheless, most existing models are limited by relatively small sample sizes, inconsistent feature selection strategies, and objectives that remain focused on lesion detection, disease classification, or volumetric assessment. In essence, they function primarily as “lesion detectors” or “volume estimators” rather than fully exploiting the deeper prognostic information embedded within routine medical images[16].

From a clinical perspective, neurosurgeons require more than accurate lesion identification. The critical challenge is to determine, immediately after hospital admission, which patients are at high risk of poor functional recovery, neurological deterioration, hematoma expansion, recurrent hemorrhage, or death, even when their baseline imaging findings appear similar[17]. Such individualized risk assessment could facilitate treatment selection, perioperative management, communication with patients and their families, rehabilitation planning, and more efficient allocation of healthcare resources. Therefore, there is a pressing need for an automated, objective, and clinically interpretable prediction framework that fully leverages routinely acquired NCCT images without requiring additional imaging sequences, specialized equipment, or complex examinations.

In this study, we developed and externally validated a fully automated machine-learning framework that integrates deep learning-based hematoma segmentation, quantitative radiomic analysis, conventional CT characteristics, and clinical variables for individualized prognostic assessment after spontaneous ICH. Using a large retrospective cohort with independent external validation, we established complementary models for predicting long-term functional outcomes and comprehensive prognostic risk stratification. In addition to evaluating predictive performance, we quantified the incremental value of radiomic features beyond conventional clinical and imaging variables. As a hypothesis-generating secondary analysis, we further explored the potential association between ambient temperature variation and disease severity as well as functional recovery. Collectively, our findings demonstrate that routinely acquired admission NCCT images contain abundant prognostic information that can improve risk stratification in patients with spontaneous ICH and support more personalized clinical decision-making.

## Methods

### Study design and participants

This is a single-center, retrospective, multi-stage imaging analysis study aimed at developing and validating a machine learning-based framework for automated hemorrhage characterization, functional network mapping, and clinical outcome prediction in spontaneous intracerebral hemorrhage (ICH).

Study participants were recruited consecutively from the clinical database of the First Affiliated Hospital of Harbin Medical University. The study population consisted of adult patients diagnosed with spontaneous ICH who underwent NCCT upon admission. The diagnosis of ICH was confirmed clinically by experienced neuroradiologists.

1. Inclusion criteria were as follows: (1) Age ≥ 18 years; (2) Confirmed spontaneous intracerebral hemorrhage by non-contrast CT (NCCT), with onset ≤ 6 hours; (3) Complete clinical records and follow-up data; (4) Baseline Glasgow Coma Scale (GCS) score 9-15; (5) Baseline National Institutes of Health Stroke Scale (NIHSS) score 0-15; (6) Baseline hematoma volume 20-40 ml; (7) CT image quality sufficient for quantitative analysis.
2. Exclusion criteria included: (1) traumatic intracranial hemorrhage; (2) hemorrhage secondary to tumors, vascular malformations, ruptured aneurysms, or hemorrhagic transformation of ischemic stroke; (3) severe liver and kidney failure; (4) a history of major neurological diseases that may interfere with prognostic assessment; (5) a history of other intracranial hemorrhages within the past 3 months; and (6) incomplete imaging or clinical data. The selection of the population fully complied with the requirement of “high matching between the target population and the intended clinical population” in ICH-E8, avoiding excessive heterogeneity that could affect the reliability of the trial results[18].

A total of 3,000 patients were initially screened. After applying the predefined eligibility criteria, 2,680 patients were included in the final analysis. Eligible patients were stratified according to baseline hematoma volume and GCS score and then randomly assigned to a training cohort (n = 1,876; 70%) and an internal validation cohort (n = 804; 30%) to ensure balanced distributions of key baseline characteristics. In addition, an independent external validation cohort (n = 196) was established from a multicenter registry using the same inclusion and exclusion criteria.

For exploratory analyses, ambient temperature at symptom onset was recorded and classified into three predefined categories according to local meteorological data matched to the date and time of stroke onset: 15°C–25°C (mild fluctuation), 25°C– 40°C (warm), and −10°C to −40°C (cold). Table 1 shows the queue characteristics of the dataset. All data were anonymized before analysis.

**Table 1:**
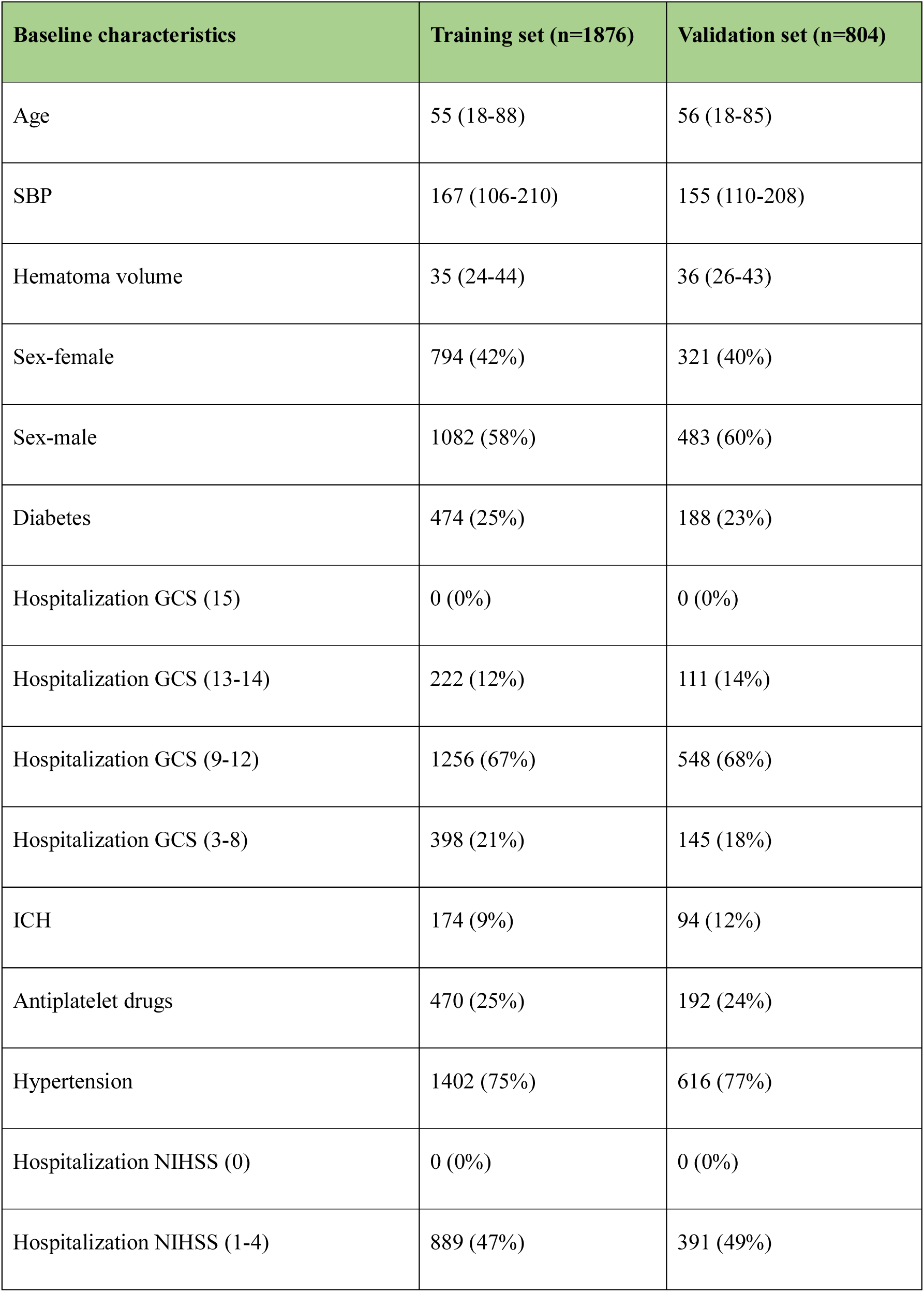
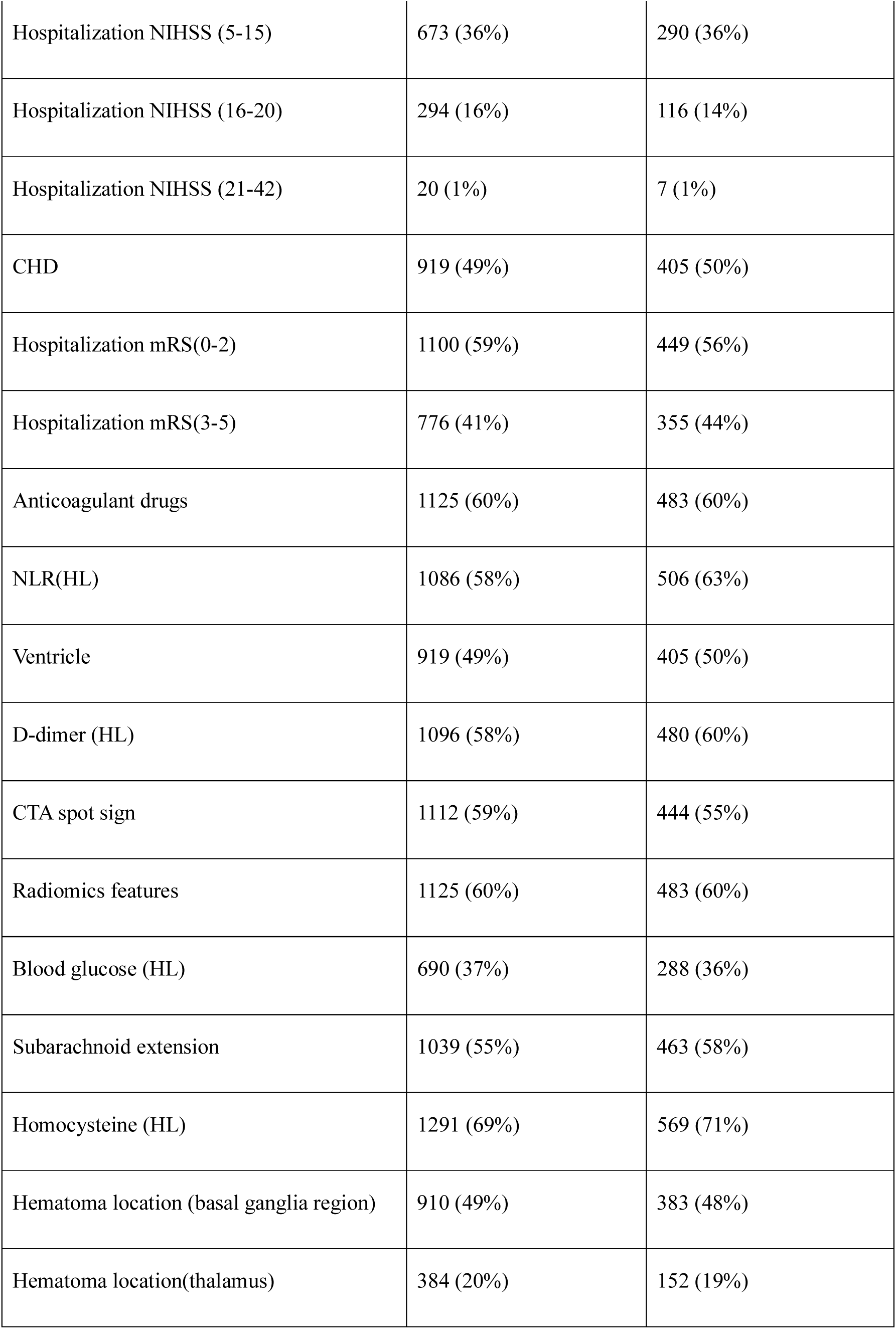
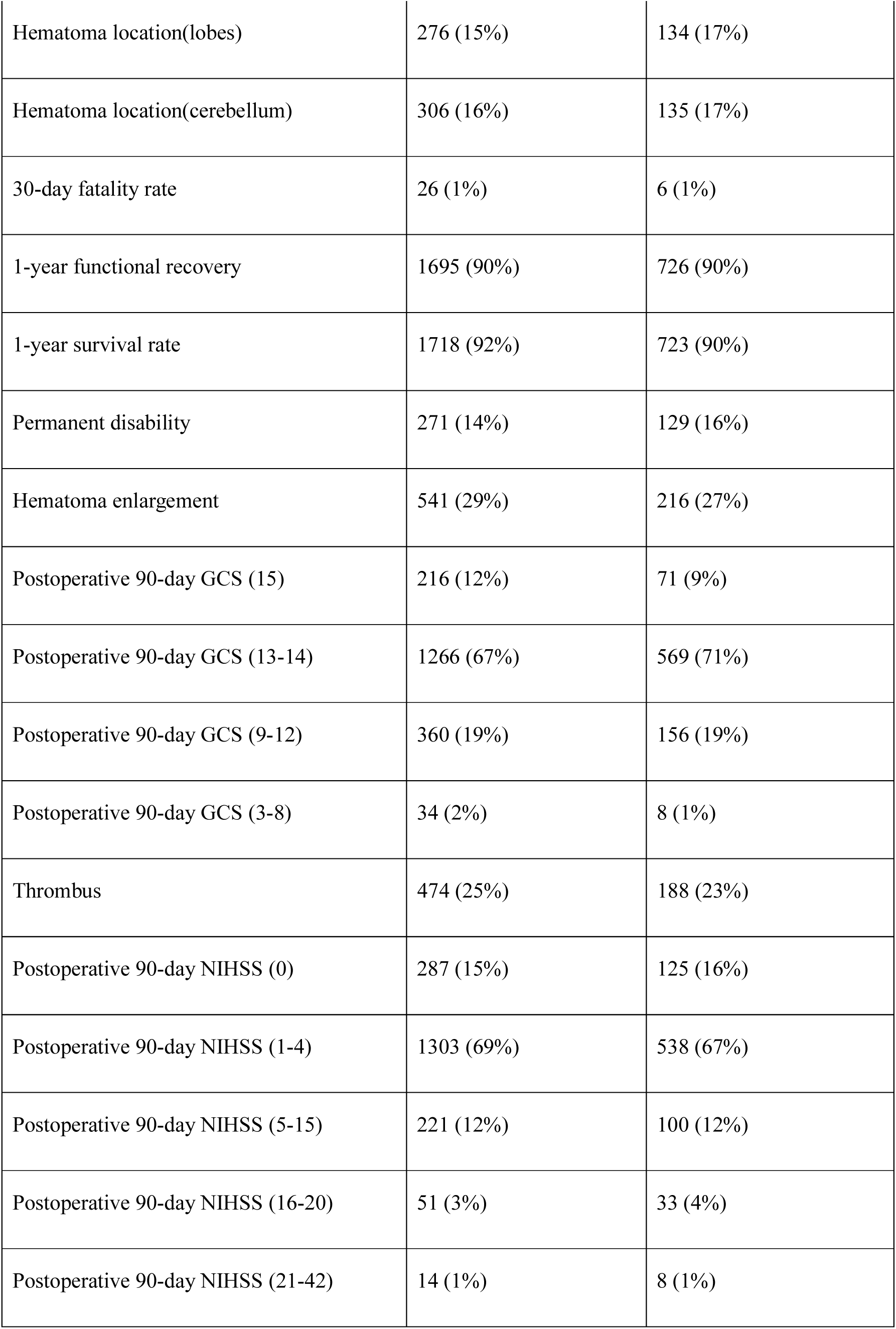
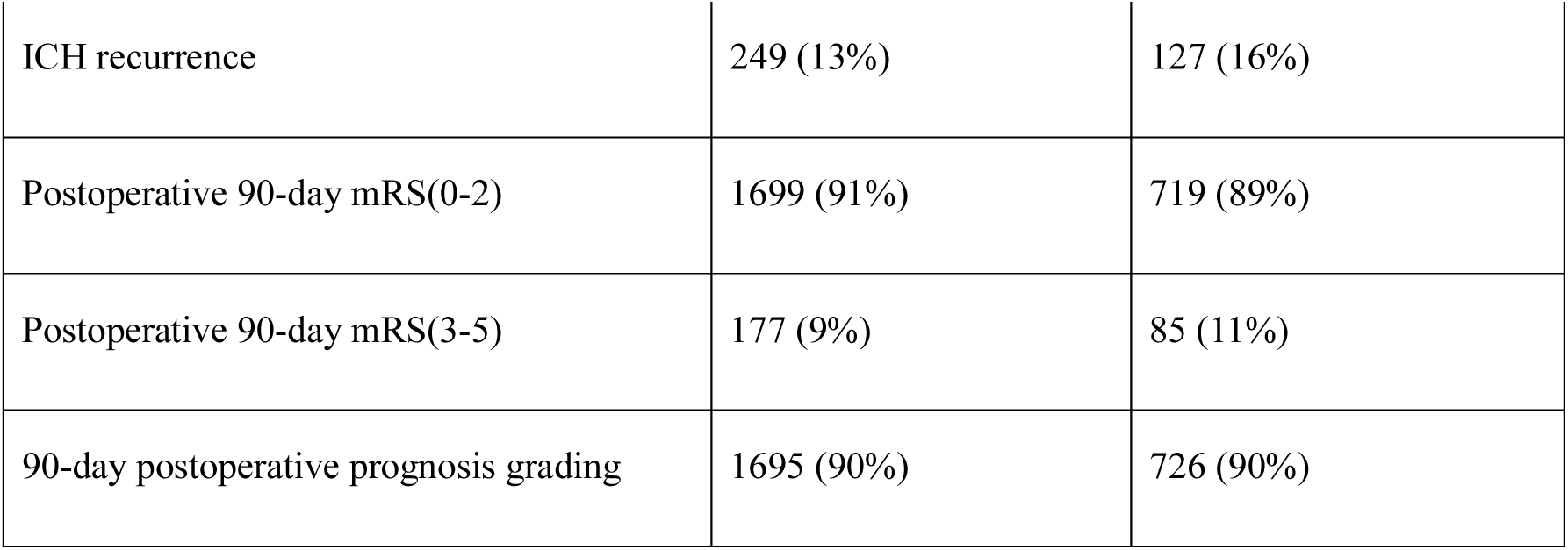
Cohort details for both datasets.

The study protocol was approved by the Ethics Committee of the First Affiliated Hospital of Harbin Medical University (2023XS19-02). The imaging and clinical database was authorized under the National Human Genetic Resources Collection License of China (No. [2023] CJ0249). All study procedures were conducted in accordance with institutional ethical guidelines and relevant regulations. As this was a retrospective study using fully de-identified data, the requirement for informed consent was waived by the ethics committee.

### Procedures

#### Image acquisition and preprocessing

All patients underwent non-contrast head CT (NCCT) according to the standard clinical imaging protocol. Before analysis, all NCCT images were preprocessed using Python 3.10.11 with the Nilearn, SimpleITK, and scikit-image libraries. First, images were resampled to an isotropic voxel spacing (1 × 1 × 1 mm³) using trilinear interpolation to ensure spatial homogeneity; intensity normalization was then performed based on a predefined Henlein unit (HU) window (0–100 HU), corresponding to brain parenchyma while excluding bone and air-related artifacts.

Skull stripping and brain extraction were subsequently performed using a hybrid approach that combined threshold-based segmentation with morphological operations, when applicable. Image quality is checked using Full-Reference (FR); the second derivative of the image is extracted using the Laplacian operator, its variance is calculated, and the largest variance is selected to ensure the consistency of the entire dataset.

To improve the visualization of hemorrhage boundaries and subtle textural details, a custom color lookup table was applied to the normalized grayscale images to generate pseudocolor images. This lookup table mapped HU values into a perceptually uniform color space, thereby enhancing the visual contrast between adjacent tissue structures. All quantitative feature extraction and model development were performed exclusively on the original grayscale NCCT images to avoid color-induced bias, whereas the pseudocolor images were used only for annotation assistance and figure presentation.

An initial hemorrhage mask was generated using binary threshold segmentation. Connected component analysis (CCA) was subsequently applied to identify contiguous foreground regions, and components containing fewer than 100 voxels were removed to eliminate image noise and isolated artifacts. For two-dimensional slice analysis, either 4-connectivity or 8-connectivity was adopted, whereas 6-, 18-, or 26-connectivity was used for three-dimensional volumetric analysis according to computational requirements. The overall image processing workflow is illustrated in Figure 1.

**Fig 1:**
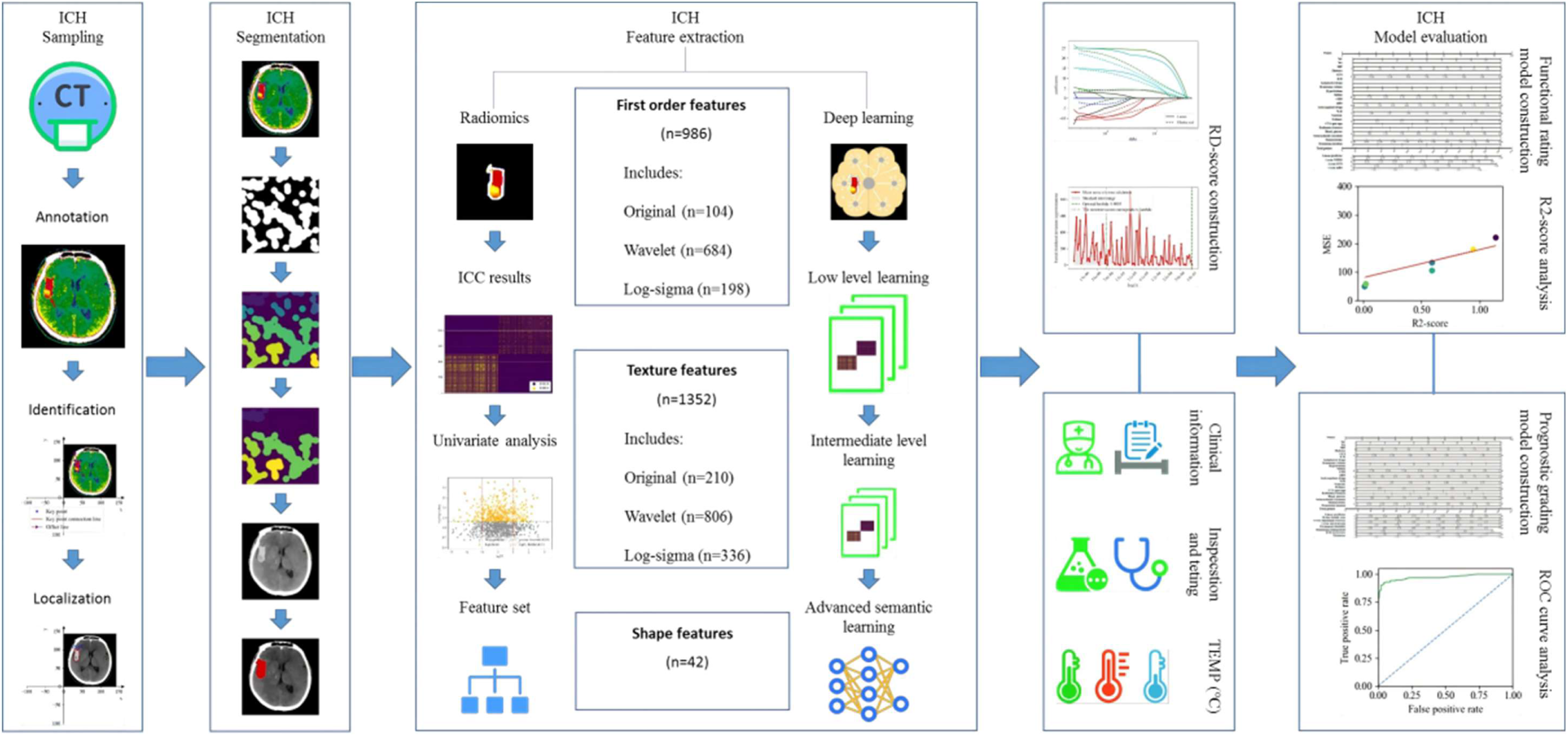
Overall workflow of the NCCT-based machine learning framework for prognostic assessment of spontaneous intracerebral hemorrhage. Schematic overview of the complete study pipeline, including patient enrollment, NCCT image preprocessing, automated hematoma segmentation, radiomics feature extraction, feature selection, machine learning model development, functional outcome prediction, prognostic stratification, and external validation.

### Hemorrhage annotation and segmentation

Intracerebral hemorrhage was initially annotated independently by two experienced neuroradiologists. Hemorrhage boundaries were manually delineated on contrast-enhanced CT images and subsequently registered to the corresponding NCCT slices using a three-dimensional rigid transformation matrix. For the initial training cohort (the first 100 patients), both the original NCCT images and their corresponding network-mapped images were generated. Expert annotations were converted into slice-wise, pixel-level label maps, establishing a one-to-one correspondence between each pixel and its class label (hemorrhage or background). For the remaining patients, pseudocolor-enhanced NCCT images were used to facilitate annotation review and quality control. Because the human visual system is substantially more sensitive to color differences than to subtle grayscale intensity variations, pseudocolor transformation subjectively expanded the perceived dynamic range and improved the visualization of fine lesion margins and morphological surface characteristics (Figure 2A, B).

**Fig 2:**
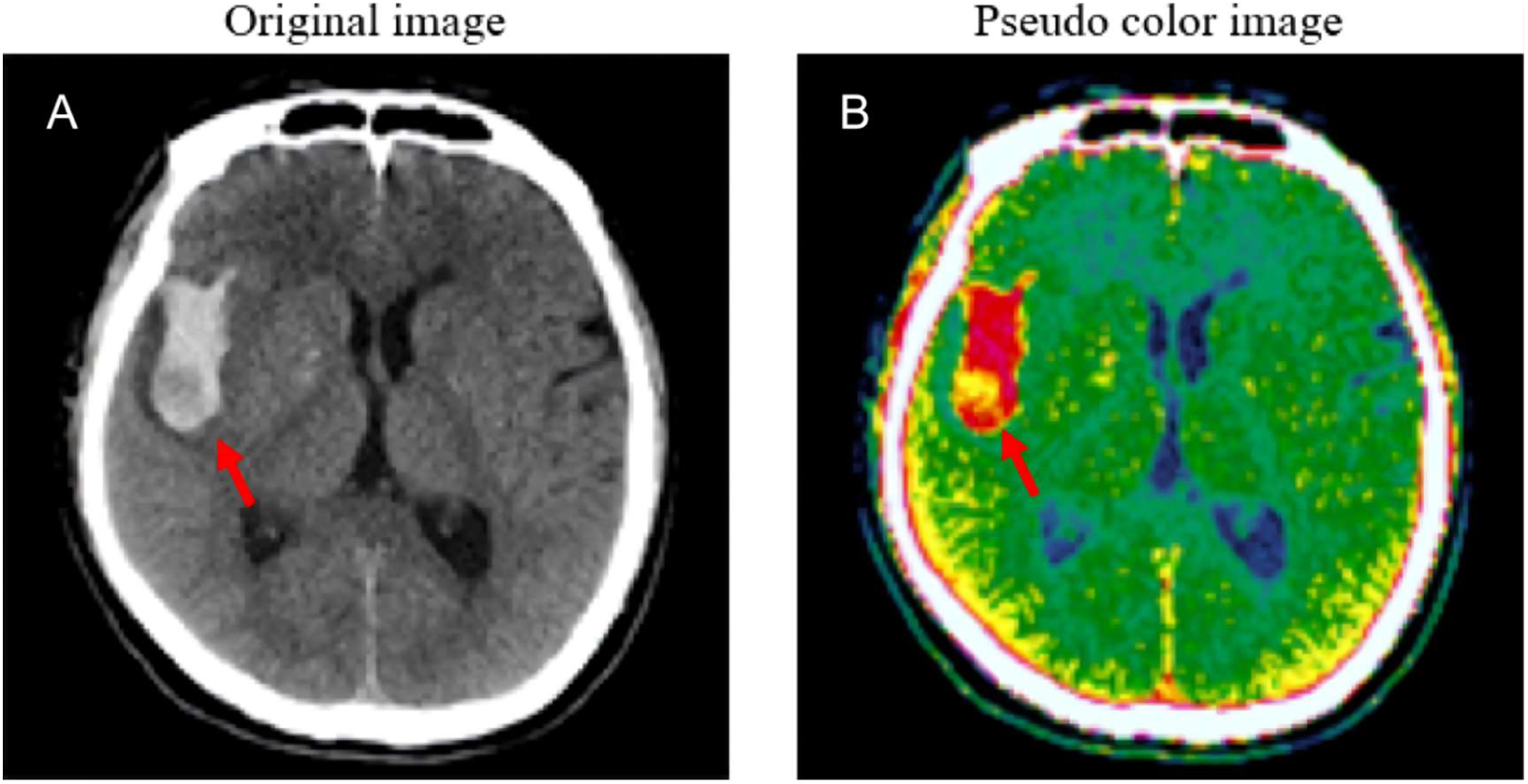
Pseudocolor-assisted visualization and annotation of intracerebral hemorrhage on NCCT images. (A) Representative original grayscale NCCT image. (B) Corresponding pseudocolor-transformed image showing enhanced visualization of hematoma boundaries and internal heterogeneity. The pseudocolor images were used to facilitate manual annotation and quality control but were not used for quantitative feature extraction or model training.

After annotation review using the pseudocolor images, an automated hemorrhage identification algorithm was deployed to systematically detect hemorrhage boundaries across all CT slices. The algorithm was based on a keypoint detection framework inspired by the Convolutional Pose Machine (CPM) and was designed to localize hemorrhage-associated vascular keypoints within the segmentation mask. Specifically, the network was trained to predict a probability distribution map for each candidate keypoint, in which the probability of a pixel belonging to a lesion boundary decreased progressively with increasing spatial distance from the corresponding ground-truth keypoint. This spatial relationship was modeled using a Gaussian kernel, and the network output consisted of probability heatmaps. The pixel with the maximum probability in each heatmap was defined as the location of the corresponding lesion-associated keypoint.

To quantitatively describe the spatial relationship between lesion boundaries and detected keypoints, we defined the shortest Euclidean distance from each detected keypoint to the lesion boundary. Let the lesion boundary be represented by the implicit curve f(x,y)=0, and let the detected keypoint be located at(dx, dy). The shortest distance from the keypoint to the lesion boundary was calculated as follows:

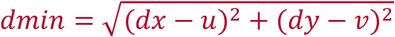

Where (u,v) is a point on the edge curve of the lesion, which must simultaneously satisfy two conditions: (1) f(u,v)=0 (i.e., the point is exactly located on the edge curve); (2) the vector (dx−u,dy−v) is parallel to the normal vector of the edge curve at that point. This derivation ensures that the identified key point-boundary distance represents the true orthogonal shortest path, providing a robust mathematical foundation for subsequent spatial contour analysis. (Figure 3A-D)

**Fig 3:**
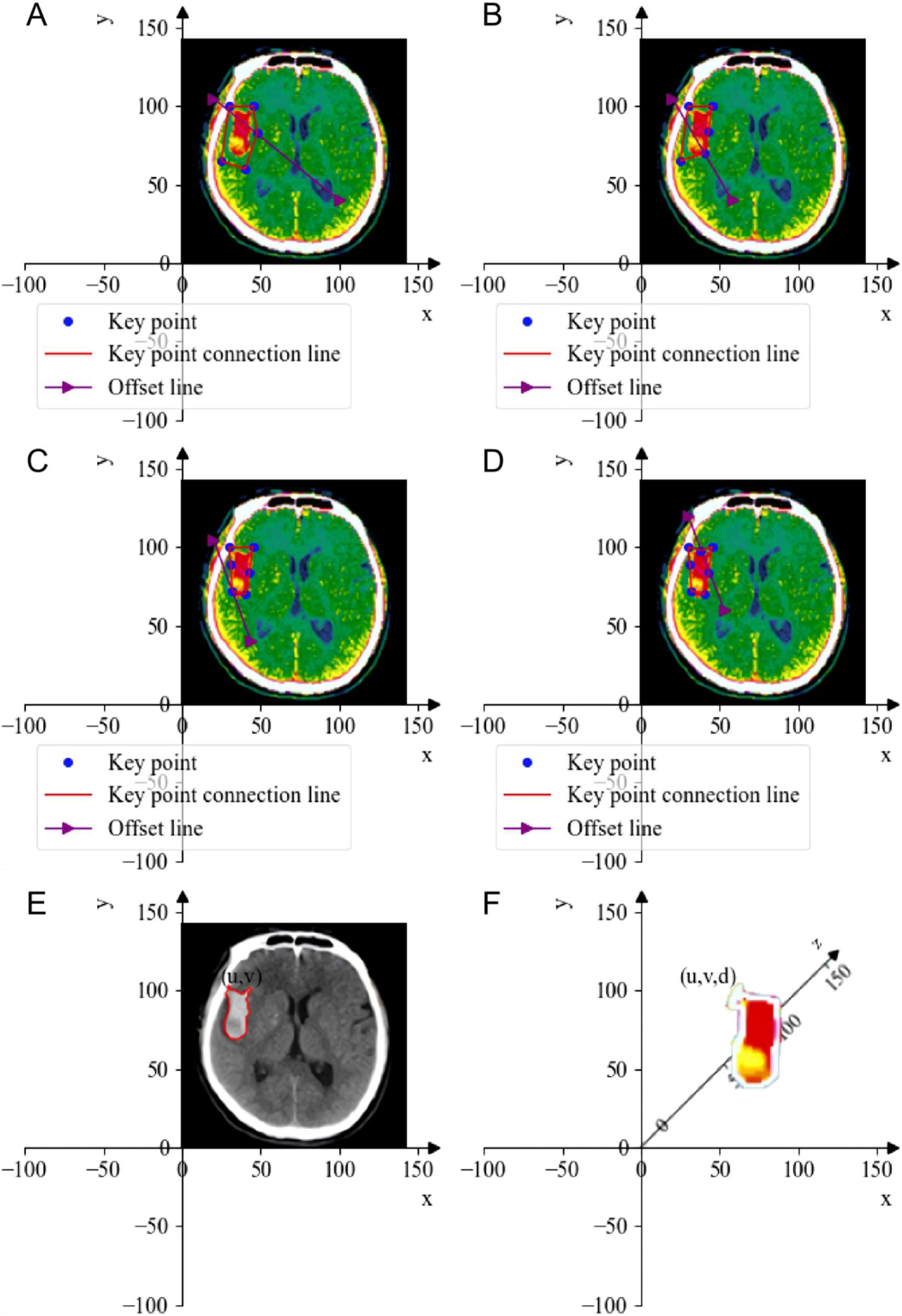
Keypoint localization and three-dimensional spatial mapping of hemorrhagic lesions. (A–D) Schematic illustration of keypoint detection and shortest orthogonal distance calculation between lesion-associated keypoints and hemorrhage boundaries. (E, F) Transformation of two-dimensional lesion coordinates into a standardized three-dimensional stereotactic coordinate system for anatomical localization and brain network mapping.

For spatial localization, the two-dimensional coordinate system was further extended into a standardized three-dimensional stereotactic coordinate space (Figure 3E, F). All segmented hemorrhage masks were registered to a common Cartesian coordinate system (X, Y, Z) with the anterior commissure serving as the origin. Two-dimensional keypoint coordinates (dx,dy) identified on each axial slice were transformed into three-dimensional volumetric coordinates by incorporating slice position information. This transformation was achieved through rigid co-registration to ensure complete spatial consistency with subsequent volume-of-interest (VOI) delineation, multimodal image registration, and image resampling procedures, thereby minimizing spatial misalignment. The resulting two-dimensional (u,v) and three-dimensional (u,v,d) coordinate sets were subsequently matched with a standardized stereotactic brain atlas, enabling each hemorrhagic lesion to be accurately assigned to its corresponding anatomical and functional brain region.

Following the initial manual annotation, a deep learning-based automated segmentation model was developed. The neural network adopted a U-Net-derived encoder-decoder architecture with skip connections. Using the manually annotated dataset as the ground truth, the model was trained for 1,600 epochs. After each training cycle, the predicted segmentation results were reviewed by experienced neuroradiologists and corrected when necessary. Through iterative training and expert refinement, the model achieved reliable localization and delineation of hemorrhagic lesions. The workflow for pseudocolor-assisted lesion segmentation and quantitative feature extraction is presented in Supplementary Figure 1.

### Radiomics feature extraction

Radiomics features were extracted from the automatically segmented 3D hemorrhagic lesion volumes. A comprehensive feature set was extracted using the PyRadiomics library, including: (1) first-order statistical features such as energy, entropy, skewness, and kurtosis; (2) shape features such as volume, sphericity, surface area-to-volume ratio, and compactness; (3) texture features derived from gray-level co-occurrence matrix (GLCM), gray-level run-length matrix (GLRLM), gray-level size-zone matrix (GLSZM), and neighborhood gray-level difference matrix (NGTDM); and wavelet transform results of all the above categories of features. A total of 1690 radiomics features were extracted for each patient.

To ensure the robustness and reproducibility of the features, the intra-group correlation coefficient (ICC) was calculated using a two-way random effects model. Two independent operators extracted features twice from a randomly selected subset of 50 patients, with a two-week interval between the two extractions. Features with intra-group and inter-group ICC > 0.75 were considered stable and retained for subsequent analysis.

### Network mapping and feature construction

The segmented hematoma was mapped onto brain functional networks defined by a standardized brain atlas using a spatial mapping function. This procedure enabled quantitative characterization of network-specific injury patterns and facilitated the investigation of their associations with clinical outcomes.

Before model training, a data generator was implemented to randomly shuffle the training samples and generate mini-batches of size *batch_size* for model optimization. In addition to the original n input features, an automated feature construction strategy was applied to generate m additional features derived from combinations and transformations of the original variables. Consequently, the final feature space consisted of n + m features.

Guided by an iterative process of model refinement and error correction, we ultimately developed an integrated machine learning framework that combined radiomic, brain network, and clinical features. The proposed model was designed to predict network alterations associated with spontaneous intracerebral hemorrhage and to estimate the probability and trajectory of functional neurological outcomes for individualized prognostic assessment.

### Model development and evaluation

Two parallel machine learning models were developed using Python 3.10.11 and the Keras framework.

Functional outcome prediction model. This model was designed to predict poor functional outcome (modified Rankin Scale [mRS] >3 at 1 year) and early neurological deterioration (END), defined as an increase of ≥4 points in the National Institutes of Health Stroke Scale (NIHSS) score or a decrease of ≥2 points in the Glasgow Coma Scale (GCS) score within 1 year. The model was implemented as a deep neural network (DNN) with multiple hidden layers. The network consisted of an input layer, followed by a Dropout layer (dropout rate = 0.25) before the first fully connected layer to reduce overfitting, three fully connected layers containing 256, 128, and 64 neurons, respectively, with rectified linear unit (ReLU) activation functions, Batch Normalization, and additional Dropout layers (dropout rate = 0.3–0.5). The output layer used a sigmoid activation function for binary classification. The model was trained using the Adam optimizer (learning rate = 1 × 10⁻⁴) with binary cross-entropy as the loss function (Figure 4A).

**Fig 4:**
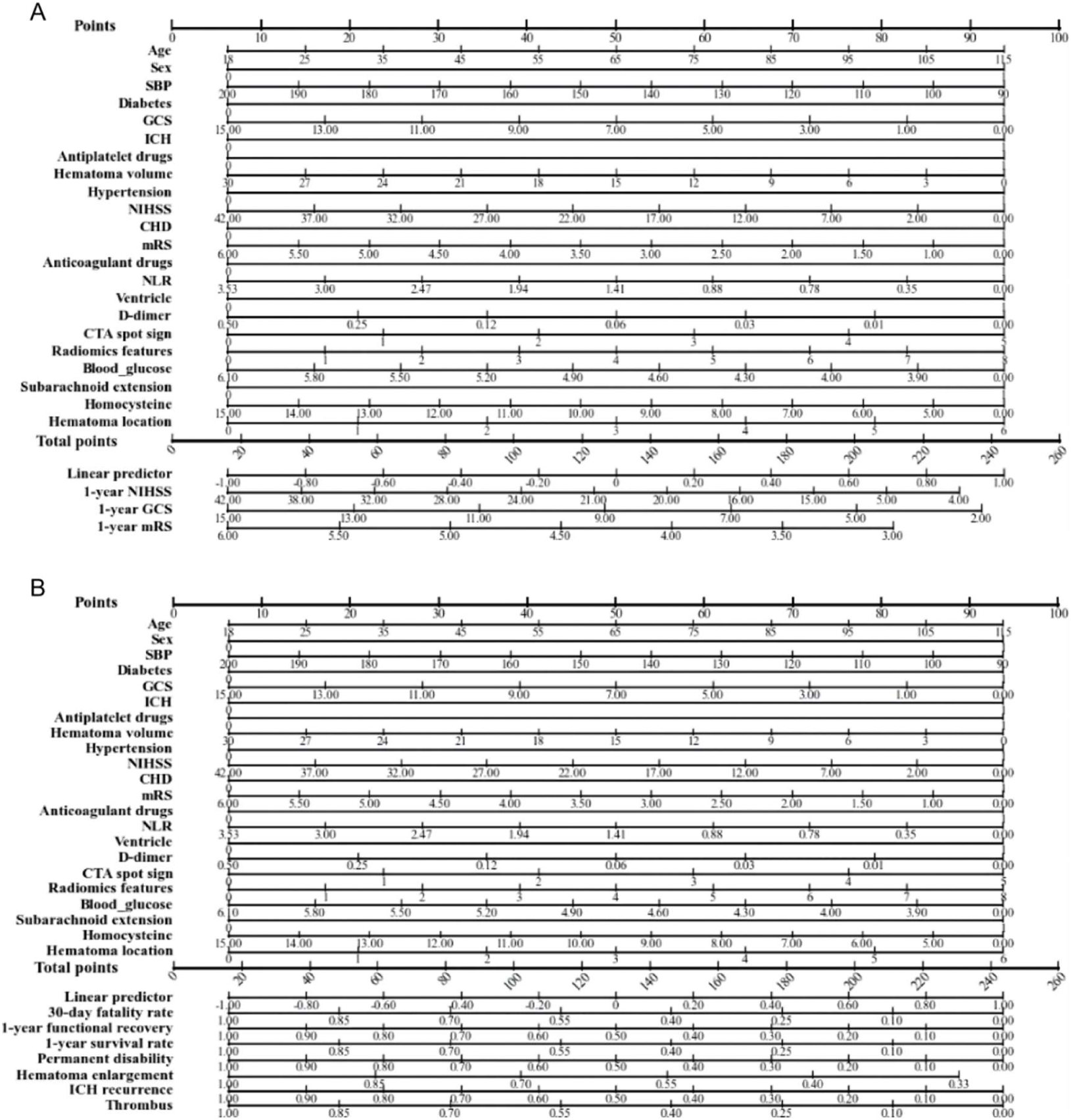
Architecture of the machine learning prediction models. (A) Deep neural network developed for prediction of long-term functional outcome and early neurological deterioration. (B) Multi-task prognostic stratification model for prediction of mortality, functional recovery, hematoma expansion, recurrent hemorrhage, thrombotic events, and other clinically relevant outcomes.

Model performance was evaluated using the mean squared error (MSE), which quantifies the average squared difference between the predicted and observed values:

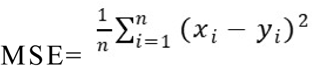

Where n is the total sample size of the validation/test set, x_i is the true result value of the i-th sample, and y_i is the model’s prediction result value for the i-th sample. A smaller MSE value indicates that the model’s prediction is closer to the true result, and the prediction accuracy is higher; conversely, a larger MSE value indicates a larger model error and poorer prediction performance. The coefficient of determination (R²-score) is used to evaluate the model’s ability to explain data variation, and the calculation formula is as follows:

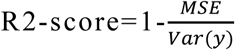

Where MSE is the mean squared error of the model, and Var(y) is the variance of the true values. The closer the R²-score is to 1, the stronger the model’s ability to explain data variation and the better its fit; R²-score = 1 indicates that the predicted values are completely consistent with the true values, and the model achieves perfect prediction; R²-score = 0 indicates that the model’s prediction performance is comparable to the baseline model that directly predicts “the mean of all samples”; R²-score < 0 indicates that the model’s prediction performance is worse than the baseline model, usually indicating severe overfitting or incorrect model selection. A general standard is: in a multi-center external validation set, R²-score > 0.7 indicates a good model fit, and R²-score > 0.5 indicates that the model has acceptable predictive ability.

Prognostic stratification model. This model was developed for comprehensive outcome stratification by simultaneously predicting multiple clinically relevant endpoints, including 30-day mortality, functional recovery at 90 days and 1 year (mRS distribution), 1-year survival, permanent disability, hematoma expansion (HE; defined as a ≥33% relative increase or an absolute increase of ≥6 mL in hematoma volume), recurrent intracerebral hemorrhage (ICH), and thrombotic adverse events. The overall network architecture was similar to that of the functional outcome prediction model, but incorporated multi-label or multi-class output heads according to the prediction task for each endpoint (Figure 4B).

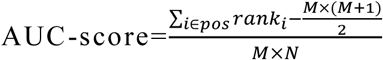

Where pos represents the set of all positive samples, M is the total number of positive samples, N is the total number of negative samples, and rank_i is the ranking of the predicted score of the i-th sample (when multiple samples have the same predicted score, the average rank of the corresponding positions is taken).

An AUC of 1.0 indicates perfect discrimination between positive and negative samples. Values between 0.85 and 0.95 indicate excellent predictive performance; values between 0.70 and 0.85 indicate acceptable discrimination with potential clinical utility; values between 0.50 and 0.70 indicate limited predictive ability, and an AUC of 0.50 indicates no discriminative capability beyond random chance.

Training strategy. Both models were trained using the training cohort (n = 1,876) with mini-batch stochastic gradient descent (batch size = 32). Model performance was evaluated on the internal validation cohort (n = 804) after each training epoch to monitor generalization performance and reduce overfitting.

External validation. The final optimized models were evaluated on an independent external validation cohort (n = 196) without further parameter tuning to assess their generalizability across different patient populations and imaging protocols.

### Statistical analysis

All statistical analyses were performed using Python 3.10.11 with standard statistical packages, including SciPy, StatsModels, scikit-learn, and lifelines. Continuous variables are presented as mean ± standard deviation (SD) for normally distributed data or as median with interquartile range (IQR) for non-normally distributed data. Categorical variables are summarized as counts and percentages.

Baseline characteristics were compared using Student’s t-test for normally distributed continuous variables and the Mann–Whitney U test for non-normally distributed variables. Categorical variables were compared using the chi-square test or Fisher’s exact test, as appropriate.

Multivariable logistic regression and Cox proportional hazards regression analyses were performed to identify independent predictors of clinical outcomes. Covariates were selected based on both clinical relevance and univariable analyses. All multivariable models were adjusted for predefined confounding factors, including age, sex, admission Glasgow Coma Scale (GCS) score, baseline hematoma volume, intraventricular extension, and time from symptom onset to imaging.

Model performance was evaluated using multiple complementary metrics. For functional outcome models with continuous or score-based endpoints, the coefficient of determination (R²) and mean squared error (MSE) were calculated. For prognostic classification models with binary outcomes, discrimination was assessed using the area under the receiver operating characteristic curve (AUC), sensitivity, specificity, positive predictive value (PPV), and negative predictive value (NPV).

Model calibration was evaluated using calibration curves and the Hosmer–Lemeshow goodness-of-fit test. Clinical utility was assessed by decision curve analysis (DCA), which quantified the net benefit across a range of clinically relevant threshold probabilities.

For the exploratory analysis of environmental temperature, patients were categorized into three groups according to the ambient temperature at symptom onset.

Multivariable regression models incorporating interaction terms between temperature group and other covariates were fitted to evaluate potential effect modification.

All statistical tests were two-sided, and a P value < 0.05 was considered statistically significant.

To ensure methodological rigor, the development and validation of the prediction models were conducted in accordance with the Transparent Reporting of a Multivariable Prediction Model for Individual Prognosis or Diagnosis (TRIPOD) Statement, while the observational components of the study adhered to the Strengthening the Reporting of Observational Studies in Epidemiology (STROBE) guidelines[19, 20].

## Results

A total of 2,680 consecutive patients with spontaneous intracerebral hemorrhage (ICH) were included in this study. Patients were randomly assigned to a training cohort (n = 1,876) and an internal validation cohort (n = 804). An independent multicenter cohort comprising 196 patients was used exclusively for external validation. Baseline demographic characteristics, vascular risk factors, admission blood pressure, neurological severity scores (Glasgow Coma Scale [GCS] and National Institutes of Health Stroke Scale [NIHSS]), hematoma volume, anatomical location, and intraventricular extension were well balanced between the training and validation cohorts, with no statistically significant differences (all P > 0.05). These findings indicate good comparability between cohorts and minimize the risk of selection bias.

### Automated lesion characterization generates reproducible radiomic biomarkers

A total of 1,690 radiomic features were extracted from each segmented ICH lesion on non-contrast computed tomography (NCCT), including first-order statistics, shape-based descriptors, and texture features derived from the gray-level co-occurrence matrix (GLCM), gray-level run-length matrix (GLRLM), gray-level size-zone matrix (GLSZM), and neighboring gray-tone difference matrix (NGTDM).

To ensure the robustness of subsequent analyses, feature reproducibility was systematically evaluated using the intraclass correlation coefficient (ICC). Features with an ICC > 0.75 were considered reproducible and retained for further analysis. The intra-group similarity heatmap (lower-left region) demonstrated high correlations among features within the same category, with distinct grid-like patterns indicating strong subgroup consistency. In contrast, the inter-group similarity heatmap (upper-right region) showed generally weak correlations across different feature categories, with only a few isolated bright-yellow signals representing minimal cross-category similarity (Figure 5A). This characteristic diagonal pattern confirmed strong within-group clustering and clear separation between different feature categories, supporting the reliability and reproducibility of the radiomic feature extraction pipeline. Of the initial 1,690 radiomic features, 1,352 (80.0%) satisfied the ICC threshold and were retained for subsequent feature selection.

**Fig 5:**
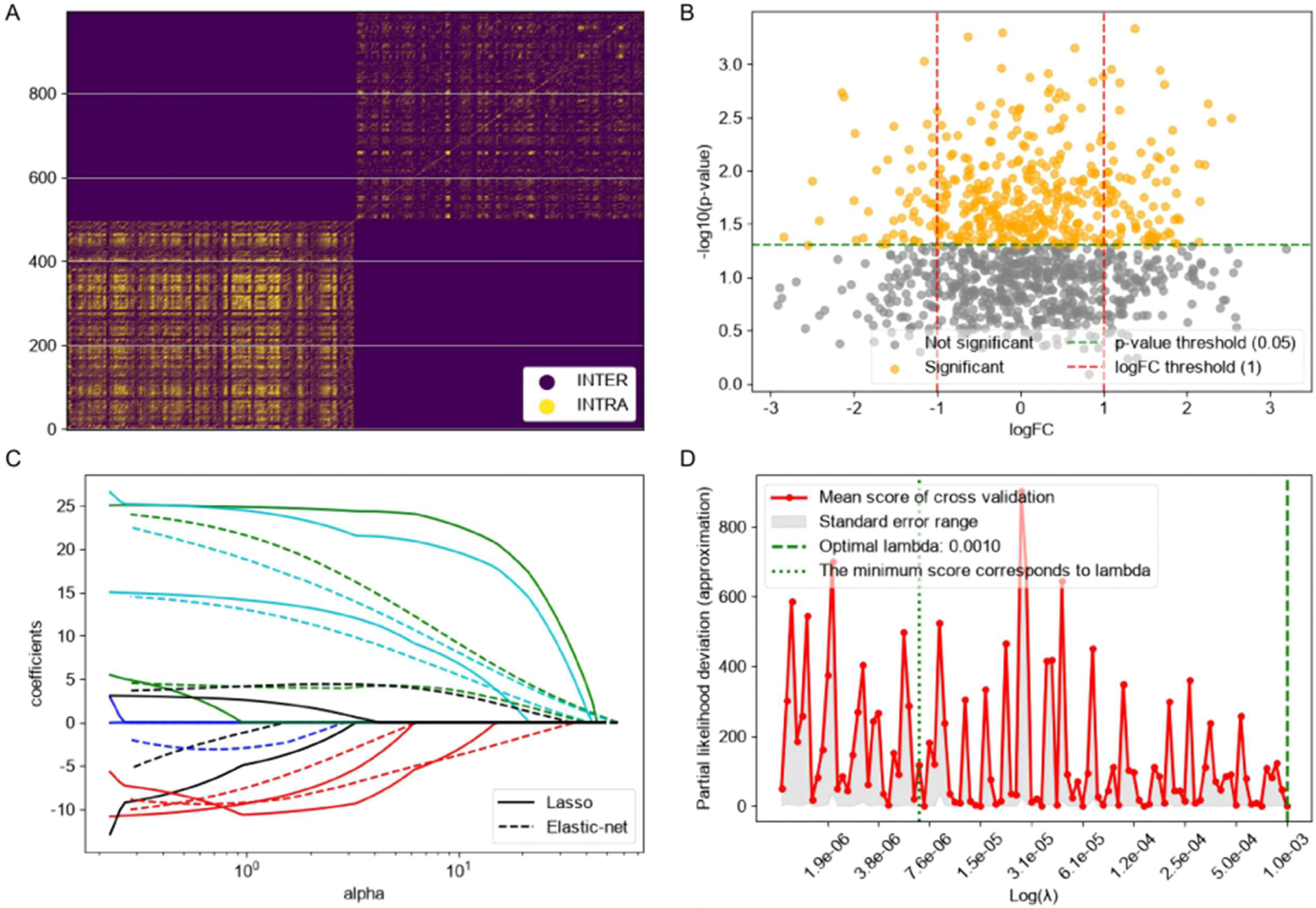
Reproducibility assessment of radiomics features. Similarity heatmap demonstrating intra- and inter-feature correlations among the extracted radiomics features. Strong within-category clustering and relatively weak between-category correlations indicate high feature reproducibility and robustness. Features with an intraclass correlation coefficient (ICC) > 0.75 were retained for subsequent analyses (A). Univariate screening of radiomics features associated with clinical outcomes. Volcano plot illustrating the results of univariable Cox regression analysis. Each point represents one radiomics feature. Features meeting the predefined statistical significance threshold were selected for subsequent LASSO regression analysis (B). LASSO-based radiomics feature selection. (B) Coefficient trajectories of radiomics features during LASSO regularization as the penalty parameter (λ) increased. (C) Ten-fold cross-validation curve showing the relationship between partial likelihood deviance and λ. The optimal λ yielded 42 radiomics features with non-zero coefficients for construction of the Rad-score (D).

### Feature selection identifies a compact radiomic signature associated with clinical outcomes

To identify the most informative features associated with functional outcomes while minimizing overfitting, a two-stage feature selection strategy was implemented.

First, univariable Cox proportional hazards regression identified 986 features with statistical significance (P < 0.05) (Figure 5B) The volcano plot showed that most features clustered around the non-significant central region, whereas subsets of significantly upregulated (right, orange) and downregulated (left, orange) features simultaneously exhibited |logFC| > 1 and statistical significance. This distribution suggests that the selected features were well balanced and demonstrated both substantial effect sizes and strong statistical confidence.

Subsequently, least absolute shrinkage and selection operator (LASSO) Cox regression with 10-fold cross-validation was performed to further optimize the feature set. At the optimal regularization parameter (λ = 0.0010), 42 features with non-zero coefficients were retained. The coefficient path plot (Figure 5 C) demonstrated that LASSO effectively shrank non-informative features toward zero while preserving the most predictive variables. The cross-validation plot of partial likelihood deviance (Figure 5 D) indicated that the selected λ achieved an optimal balance between model performance and generalizability by minimizing deviance while maintaining regularization. A radiomics score (Rad-score) was then calculated for each patient using the selected 42 features according to the following equation: Rad-score = β₀ + ΣβₙFₙ, where βₙ represents the regression coefficient of the nth radiomic feature, and Fₙ denotes its corresponding feature value.

### Radiomics provides substantial incremental value for functional outcome prediction

An integrated prognostic model was developed by combining demographic characteristics, clinical variables, conventional radiological features, and the derived Rad-score. The model was trained to predict poor functional outcome, defined as a modified Rankin Scale (mRS) score >3 at 1 year, as well as early neurological deterioration (END), defined as an increase of ≥4 points in the NIHSS score or a decrease of ≥2 points in the GCS score within 1 year.

Model performance was evaluated by the coefficient of determination (R²) across successive stages of model development. The clinical model alone (Figure 6A) demonstrated limited explanatory power (MSE = 387, R²= 0.35), indicating that clinical variables accounted for only a modest proportion of outcome variability. Incorporation of conventional radiological features (Figure 6B) improved model performance (MSE = 263, R²= 0.56), suggesting that imaging findings contributed additional prognostic information. Notably, the addition of radiomic and network-derived features (Figure 6C) substantially enhanced predictive performance (MSE = 106, R²= 0.72), exceeding the clinically meaningful threshold of R²= 0.70. In the fully optimized model incorporating all feature categories (Figure 6D), the R² further increased to 0.93 with a corresponding MSE of 61, demonstrating excellent explanatory capability. The progressive improvement in R² from 0.35 to 0.93 highlights the substantial incremental value provided by radiomic and network-derived features beyond conventional clinical and radiological markers.

**Fig 6:**
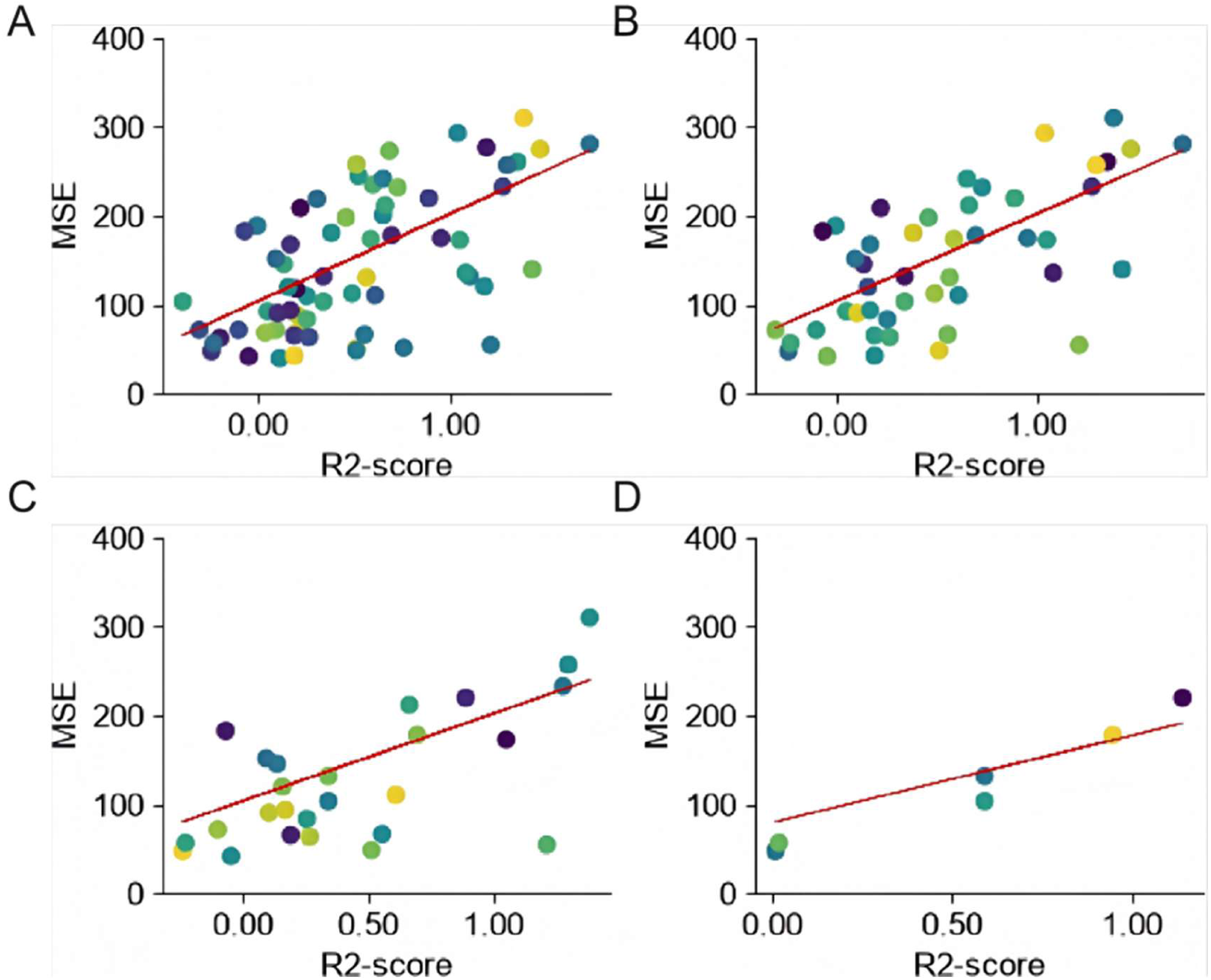
Incremental predictive performance of the model for functional outcome prediction. The scatter plots show the correlation between predicted and observed functional outcomes (mRS at 1 year), with the R² and MSE values for each sequential model stage. (A) The model based solely on clinical variables. (B) The model incorporating conventional radiological features. (C) The model after the addition of radiomic and network-derived features. (D) The fully optimized integrated model, demonstrating a substantial improvement in the coefficient of determination (R² from 0.35 to 0.93) and a significant reduction in mean squared error (MSE from 387 to 61), highlighting the incremental value provided by radiomic features.

Calibration curve analysis demonstrated excellent agreement between predicted and observed probabilities, with the calibration curve closely approximating the ideal diagonal reference line (Figure 7A). Decision curve analysis further confirmed the clinical utility of the integrated model, showing consistently greater net benefit than both the “treat-all” and “treat-none” strategies across clinically relevant threshold probabilities (Figure 7B).

**Fig 7:**
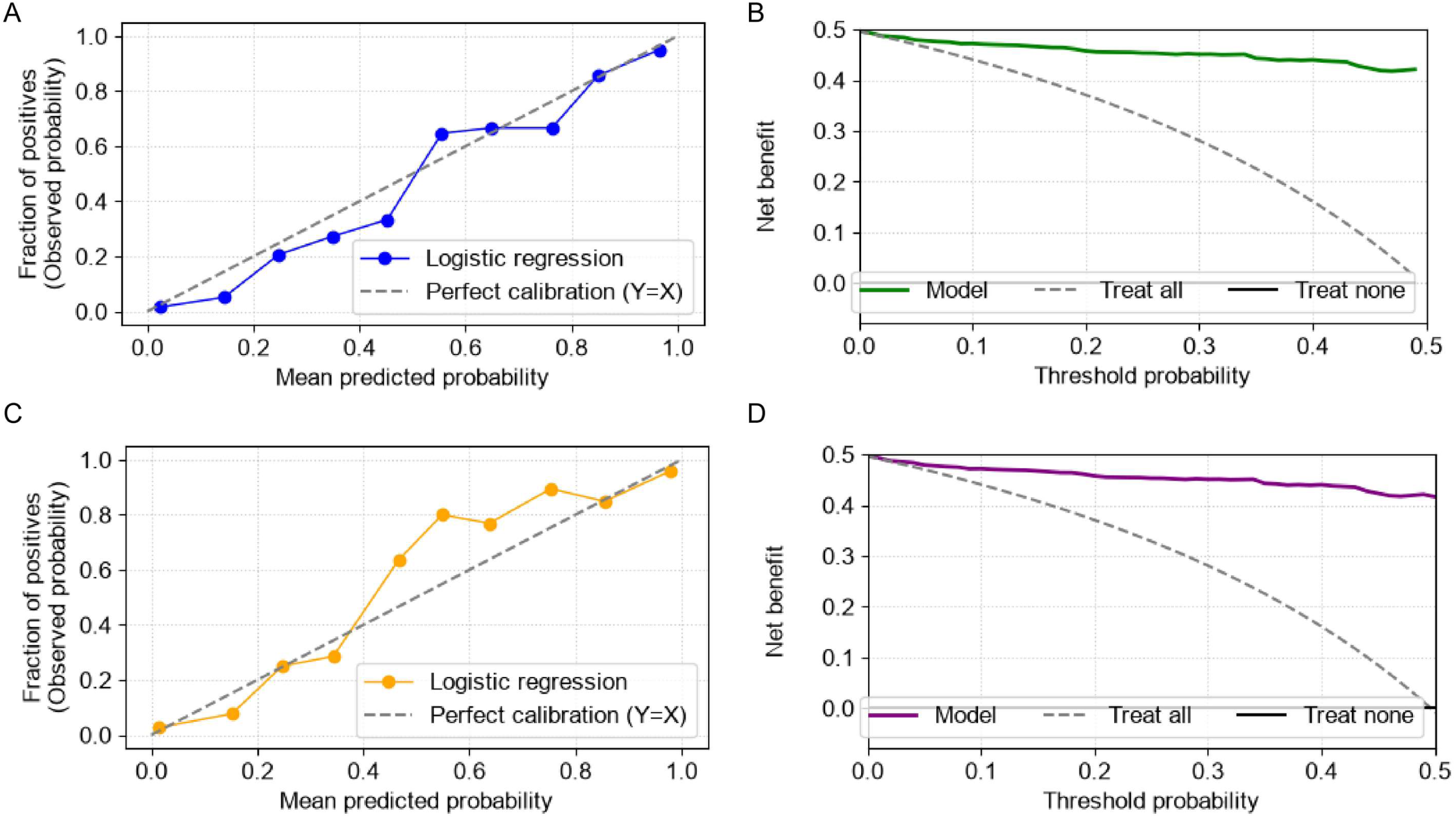
Calibration and clinical utility of the integrated models. (A-B) Performance of the Functional Outcome Prediction Model. (A) Calibration curve showing excellent agreement between the predicted probability of poor functional outcome and the observed frequency across the entire probability spectrum, closely approximating the ideal diagonal reference line. (B) Decision curve analysis (DCA) demonstrating the net clinical benefit of the integrated model across a range of threshold probabilities. The model’s net benefit consistently exceeds both the “treat-all” and “treat-none” strategies. (C-D) Performance of the Prognostic Stratification Model. (C) Calibration curve showing close alignment between predicted and observed event rates. (D) DCA curve confirming the model’s clinical utility with a consistently higher net benefit across clinically relevant decision thresholds.

### The integrated model achieves robust prognostic stratification in independent cohorts

The discriminative performance of the prognostic model was assessed using the area under the receiver operating characteristic curve (AUC). The model based solely on clinical variables (Figure 8A) achieved an AUC of 0.59, indicating discrimination only marginally better than chance. After incorporation of conventional radiological features (Figure 8B), the AUC increased to 0.71, reaching an acceptable level for clinical prediction. The addition of radiomic features (Figure 8C) further improved discrimination, yielding an AUC of 0.84. Finally, the fully integrated model incorporating clinical, radiological, radiomic, and network-derived features (Figure 8D) achieved an AUC of 0.95, demonstrating excellent discriminative performance.

**Fig 8:**
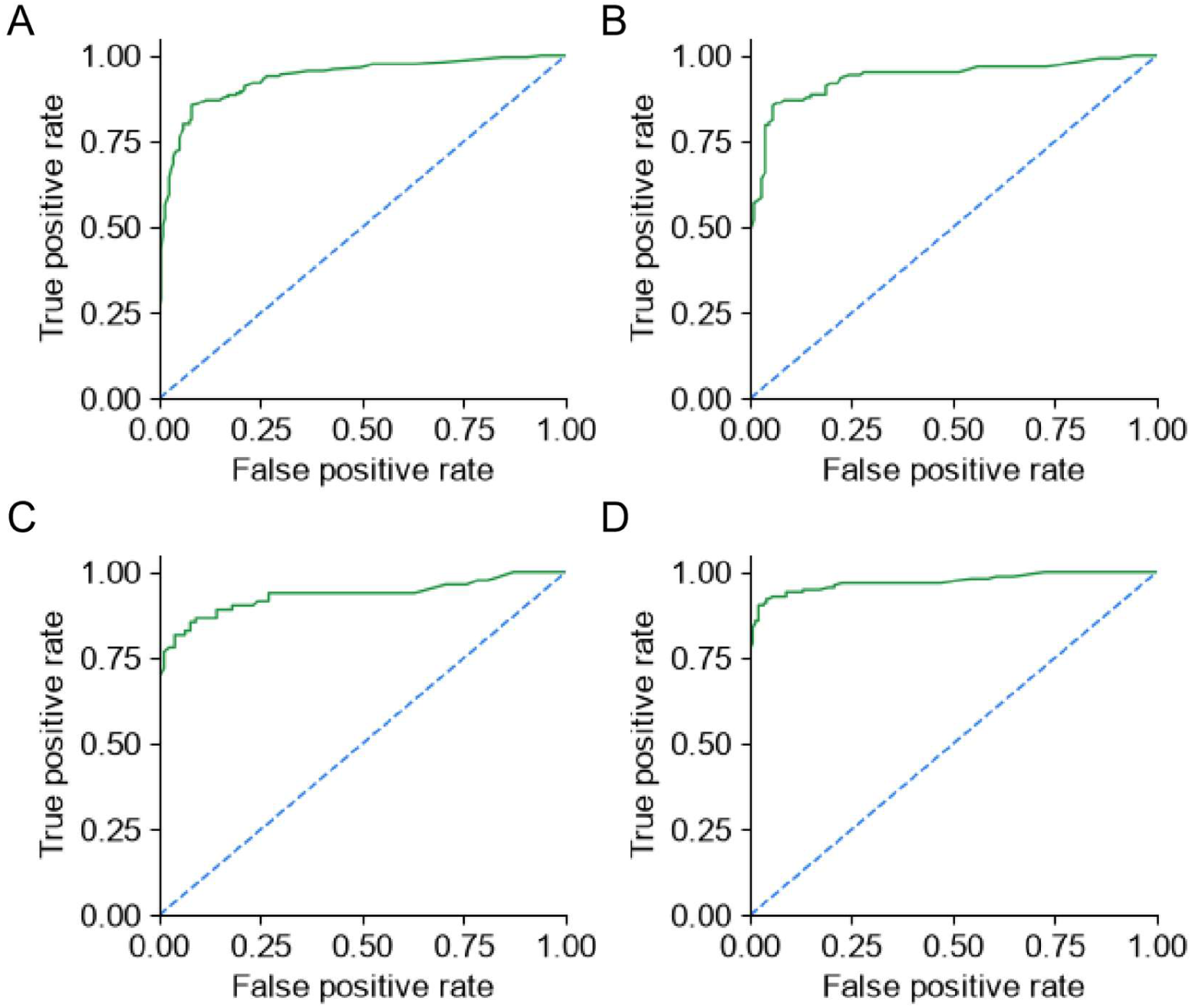
Receiver operating characteristic (ROC) curves for the prognostic stratification model. The ROC curves and corresponding area under the curve (AUC) values illustrate the discriminative performance of the model for predicting poor prognosis across its sequential stages in the training cohort. (A) The clinical-only model (AUC = 0.59). (B) The model after adding conventional radiological features (AUC = 0.71). (C) The model after further incorporating radiomic features (AUC = 0.84). (D) The fully integrated model with all features (AUC = 0.95). The model maintained robust performance in the independent external validation cohort (AUC = 0.86), confirming its generalizability.

To further examine the model’s generalization ability on data not used in its training, we employed an independent external validation cohort, CQ196. This cohort included 196 patients, of whom 137 were positive and 59 were negative. This cohort was not involved in model training or internal validation. In this completely independent real-world cohort, the model maintained stable discrimination and quantitative performance overall, with sensitivity of 0.90–0.91, specificity of 0.94–0.95, R² of 0.89–0.93, and AUC of 0.93–0.95 (Table 2). Notably, the model exhibited high and relatively stable performance even across different hematoma volumes: for microbleeds <5 mL, the AUC was 0.93; for hematomas of 5–10 mL, 10–15 mL, 15–30 mL, 30–40 mL, and >40 mL, the AUCs were 0.92, 0.95, 0.98, 0.95, and 0.96, respectively. The 15–30 mL hematoma subgroup showed the highest discriminative ability (AUC=0.98, 95% CI 0.95–1.00), and even in the most challenging micro-bleeding subgroup, the model maintained high sensitivity and specificity, suggesting good robustness to different hematoma loads. For large hematomas (>40 mL), the model sensitivity reached 0.97–0.98, further demonstrating its strong ability to identify significant hemorrhagic lesions.

**Table 2:**
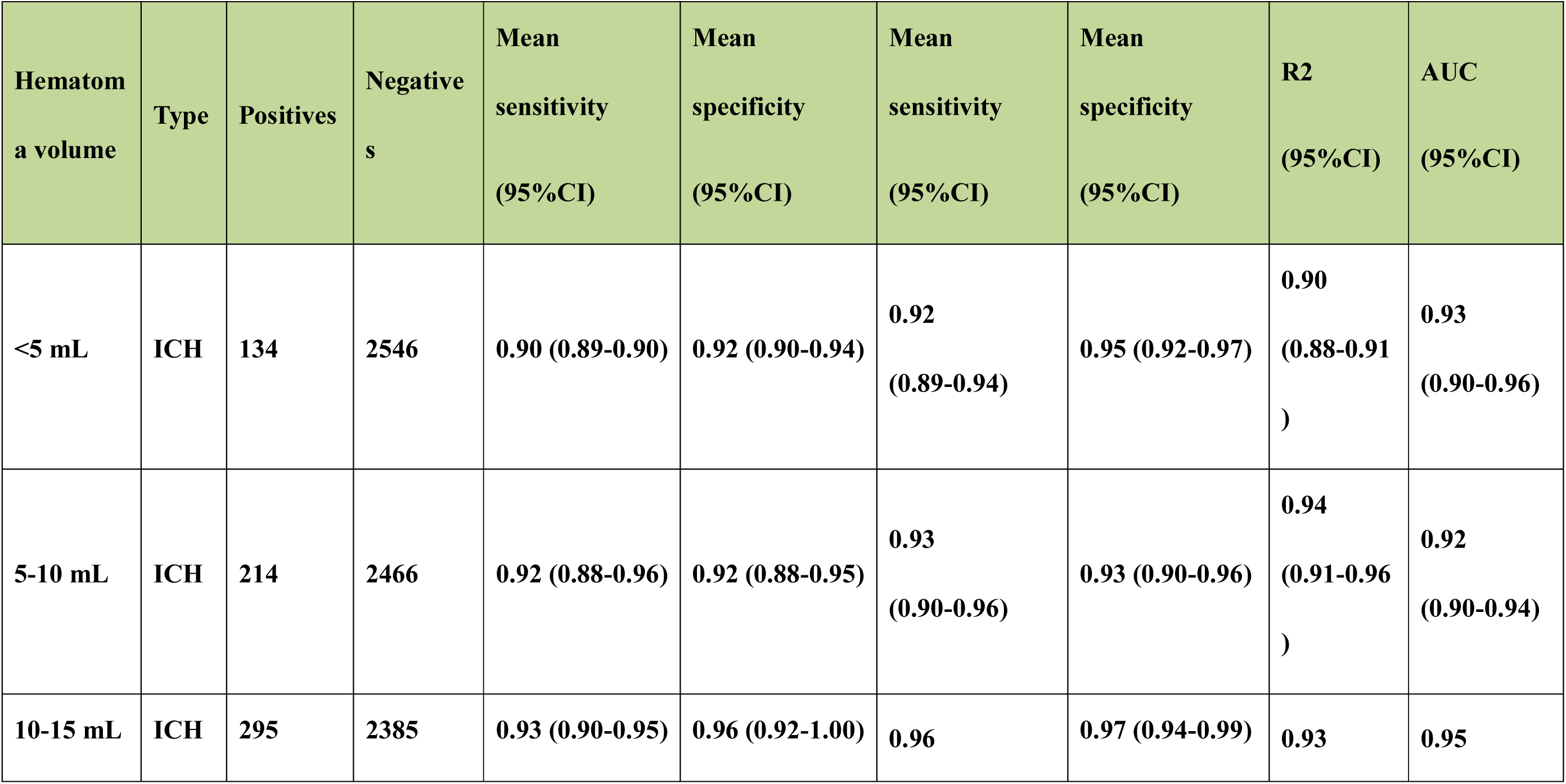
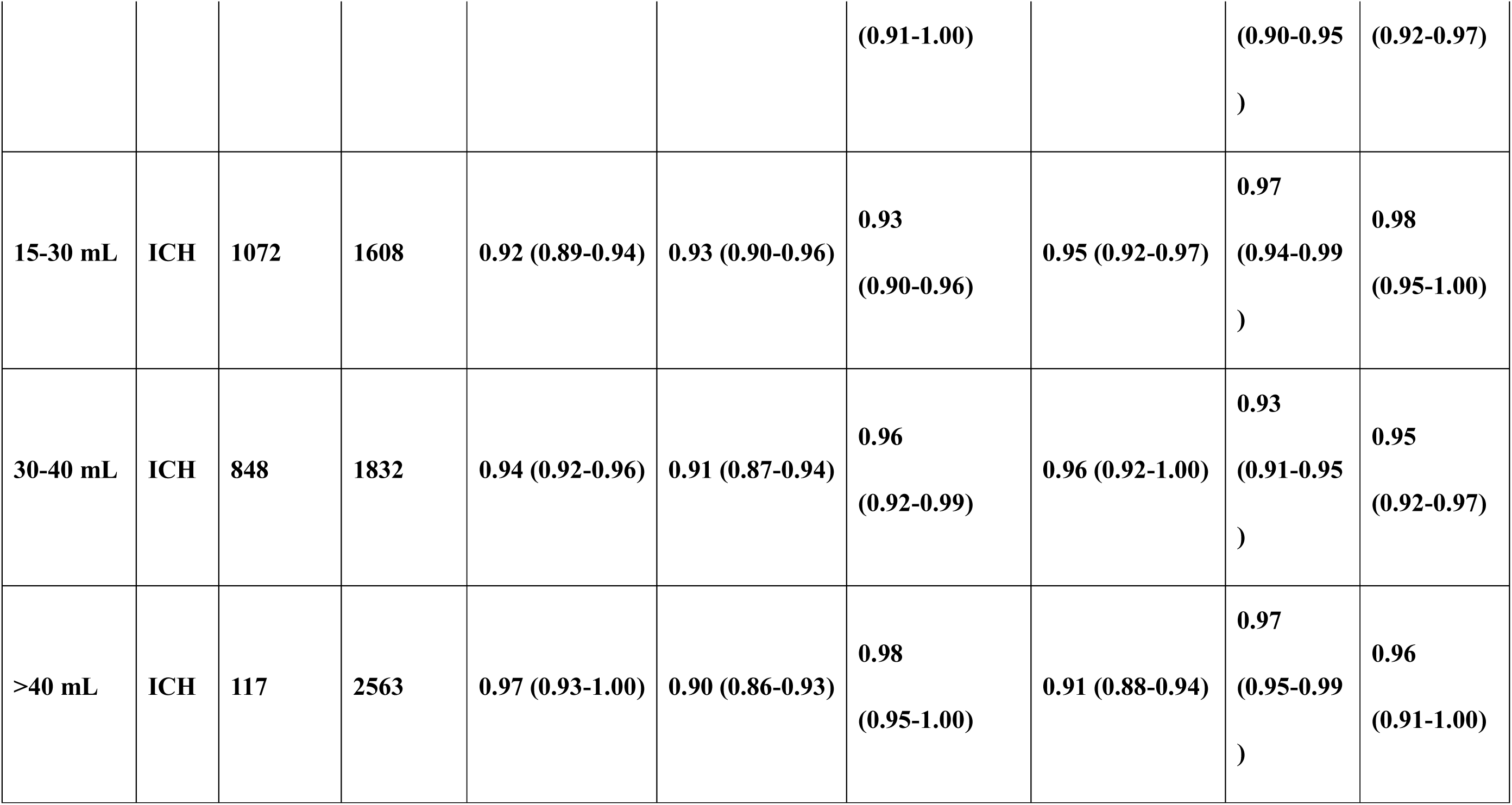
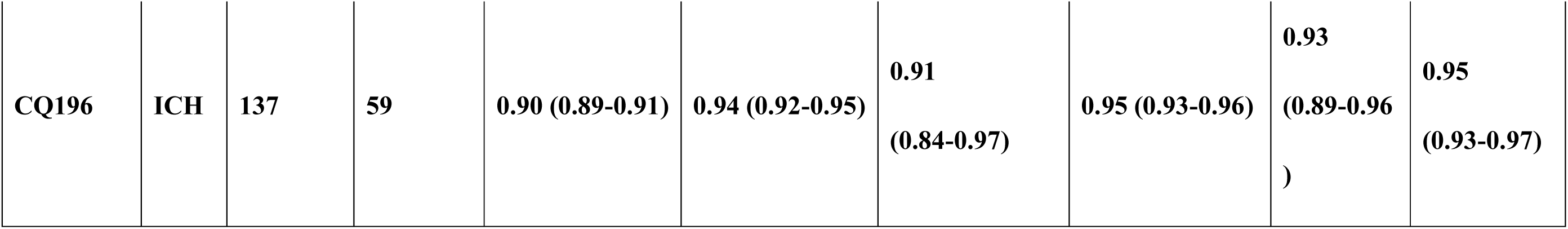
Test results of the external validation set (CQ196).

Building upon the validation of the independent imaging and hematoma quantification performance, we further evaluated the generalization ability of the complete prognostic model in the CQ196 cohort. Compared to the internal validation results, the model still achieved an AUC of 0.86 in a completely independent external cohort, indicating good prognostic discrimination in data not involved in model development.

We further used calibration curves to assess the consistency between the model’s predicted probabilities and actual observations. The results showed that the model’s predicted probabilities maintained high consistency with actual observed incidence rates across the entire risk range, with only a small deviation from the ideal diagonal (Figure 8C), suggesting that the model not only has good discrimination ability but also good probability calibration performance. Decision curve analysis (DCA) further evaluated the potential clinical benefit of the model at different clinical risk thresholds. The results showed that within the clinically meaningful threshold probability range, the net benefit of the complete model was significantly higher than that of the “all intervention” and “no intervention” reference strategies (Figure 8D), suggesting that the model’s predictive results have potential clinical decision-making value.

## Discussion

In this study, we developed and externally validated a non-contrast computed tomography (NCCT)-based clinical decision support system (CDSS) for patients with intracerebral hemorrhage (ICH). By integrating deep learning-based hematoma segmentation, quantitative radiomics, conventional CT imaging features, and clinical variables, the proposed framework demonstrated excellent performance in predicting long-term functional outcomes and comprehensive prognostic stratification. More importantly, our sequential modeling strategy showed that radiomics provided substantial incremental predictive value beyond conventional clinical and imaging variables. Rather than focusing solely on algorithm development, this study was designed to address a clinically relevant question: how routine admission NCCT can be transformed into an objective and individualized decision-support tool. Our framework achieved fully automated hematoma segmentation without manual intervention and enabled robust extraction of high-dimensional quantitative imaging biomarkers from routine NCCT. By combining radiomics with conventional clinical and imaging features, the model achieved excellent discrimination, calibration, and clinical utility while maintaining robust performance in an independent external validation cohort.

Recent years have witnessed remarkable advances in deep learning for the detection and segmentation of intracranial hemorrhage on head CT[21]. Monteiro et al. developed a convolutional neural network using the multicenter CENTER-TBI dataset for voxel-wise segmentation of four traumatic brain injury lesion types, including intraparenchymal hematoma, extra-axial hematoma, intraventricular hemorrhage, and peri-lesional edema. Their model achieved an area under the receiver operating characteristic curve (AUC) of 0.83 for intracranial hemorrhage detection in the external dataset[22]. However, their framework primarily focused on lesion quantification and was not extended to prognostic prediction or clinical decision support. In the present study, pseudo-color enhancement was introduced during image preprocessing to simplify the convolutional neural network pipeline while improving lesion boundary recognition, particularly for low-contrast hematomas. Beyond accurate hematoma segmentation, the extracted radiomic features were directly linked to clinically meaningful endpoints through machine deep learning models, achieving an R² of 0.93 for functional outcome prediction and an AUC of 0.95 for prognostic classification.

Machine learning has also shown considerable promise in predicting outcomes after ICH[23]. Nevertheless, most previous studies have primarily pursued optimal predictive performance, with relatively limited emphasis on clinical translation[24]. Yuan et al. developed a gradient boosting decision tree (GBDT) model based on 215 patients with spontaneous ICH to predict the 180-day Glasgow Outcome Scale (GOS), achieving an AUC of 0.865 in internal validation but declining to 0.785 in external validation[25]. A multicenter study including 380 patients reported an AUC of 0.969 using an extreme gradient boosting (XGBoost) model to predict 3-month functional outcomes[26]. Pei et al. constructed a combined clinical-radiomics model using 161 patients and achieved an AUC of 0.915 in the validation cohort[27]. Although these studies demonstrate the technical feasibility of machine learning for ICH prognosis, most models remain “black boxes,” making it difficult for clinicians to understand the basis of individual predictions. In addition, prognostic outputs are typically presented as static binary classifications, which fail to capture the dynamic progression of disease and therefore provide limited guidance for individualized clinical decision-making.

The CDSS framework used in this study is consistent with the recent research trend of interpretability models using the SHAP (Shapley Additive Explanations) method[25, 28, 29]. Through visualization of the LASSO feature selection process, coefficient trajectories, and the stepwise improvement in model performance across sequential modeling stages, clinicians can intuitively understand the contribution of different feature groups to the final prediction. In addition, our findings are consistent with those of Liang et al., who suggested that anatomical location features are more robust and interpretable than radiomic features alone. Accordingly, our framework integrates clinical variables, conventional imaging features, and radiomic biomarkers, while quantitatively demonstrating the incremental contribution of each feature category through progressive model improvement across four stages (R²: 0.35→0.56→0.72→0.93; AUC: 0.59→0.71→0.84→0.95). Furthermore, by calculating the Shapley value, the fair contribution of each feature to the model’s prediction results is quantified, decomposing the prediction of the complex black-box model into the independent contribution value of each feature.

An exploratory analysis further identified a potentially important environmental factor associated with ICH severity and recovery: daily ambient temperature fluctuation on the day of symptom onset. The relationship between environmental temperature and ICH incidence has been well documented in epidemiological studies. Multiple reports have consistently demonstrated a higher incidence of ICH during colder seasons. A large cohort study with data spanning 20 years showed that the incidence of cerebral hemorrhage has a seasonal peak in winter[30]. A study based on INTERACT2 showed that the risk of non-invasive ICH is significantly increased during winter (ambient temperature <10°C)[31]. Our exploratory findings are generally consistent with these observations. Across three predefined temperature ranges (15–25°C, 25– 40°C, and −10 to −40°C), significant differences were observed in lesion extent, hematoma absorption, blood pressure responses, and clinical outcomes. During the acute phase, patients in Group C (−10 to −40°C) exhibited the largest lesion volumes and the highest imaging intensity, indicating the greatest initial injury burden (Supplementary Figure 2). During the subacute phase, Group C also showed the slowest hematoma absorption, suggesting delayed clearance of injured tissue under conditions of marked temperature fluctuation. Blood pressure monitoring (Supplementary Figure 3) further suggested a possible mechanistic link. Patients in Group C experienced the greatest acute systolic blood pressure elevation and the slowest blood pressure recovery during the subacute phase. Regression analyses showed similar slopes among the three temperature groups but a clear gradient in the estimated probability of ICH occurrence: P (−10 to −40°C) > P (25–40°C) > P (15– 25°C) (Supplementary Figure 4). These findings suggest that the magnitude of temperature fluctuation, rather than exposure to low or high temperature alone, may be an important factor influencing blood pressure variability, ICH severity, and the likelihood of spontaneous ICH. Nevertheless, these observations should be interpreted cautiously because the temperature data were derived from meteorological records rather than individual-level exposure, and the association between temperature and ICH outcomes may vary across geographical regions. Prospective epidemiological studies integrating environmental and imaging data are therefore required to validate these findings.

Several limitations should be acknowledged. First, the retrospective single-center design may have introduced selection bias. Although the model demonstrated encouraging performance in an independent external validation cohort (n = 196), larger prospective multicenter studies are still required. Second, the impact of the proposed framework on clinical workflow and reporting efficiency has not yet been evaluated in real-world practice. Previous work by O’Neill et al. has shown that different methods of AI implementation, such as worklist prioritization versus passive notification, can substantially influence clinical efficiency. Therefore, prospective deployment studies are needed to assess the practical clinical value of our system. Finally, only patients with supratentorial ICH were included in this study, and the generalizability of the proposed framework to infratentorial ICH remains to be established.

In conclusion, we developed an automated and interpretable NCCT-based clinical decision support system that integrates deep learning-based hematoma segmentation, radiomic feature extraction, machine learning, and cohort theory-based prioritization into a fully automated workflow from lesion detection to prognostic prediction. Both the functional outcome prediction model and the prognostic classification model demonstrated excellent performance in internal and external validation cohorts. In addition, our exploratory analysis identified ambient temperature fluctuation as a potential modifier of ICH severity and prognosis, providing new insights into seasonal prevention strategies. Future work will focus on prospective multicenter validation, evaluation of clinical workflow integration in real-world settings, and further investigation of the biological mechanisms linking environmental temperature variation with ICH.

## Author Contributions

C.L. and Q.S. contributed equally to this work. C.L. and Q.S. conceived and designed the study, performed the statistical analyses, and drafted the main manuscript text. Y.L. and C.Y. developed the deep learning segmentation and radiomics analysis pipeline and prepared Figures 1–3. H.X. and Y.X. performed model validation and data curation. C.Y. and J.Z. collected and verified the clinical data. Y.L. and G.Y. conceived and supervised the study, acquired funding, and critically revised the manuscript. All authors reviewed and approved the final manuscript.

## Data sharing

The de-identified imaging and clinical data supporting the findings of this study are not publicly available because they contain protected patient information and are subject to institutional and national regulations governing human genetic resources. Access to the data may be granted by the corresponding author upon reasonable request, subject to approval by the Ethics Committee of the First Affiliated Hospital of Harbin Medical University and compliance with the National Human Genetic Resources Administration of China, institutional data-sharing policies, and applicable legal and ethical requirements.

## Acknowledgments

We are grateful to the neuroradiologists and neurosurgical staff at the First Affiliated Hospital of Harbin Medical University for their assistance with data collection and image annotation. We also acknowledge the contributions of the collaborating centers that provided the external validation cohort.

---

**Sup Fig 1:**
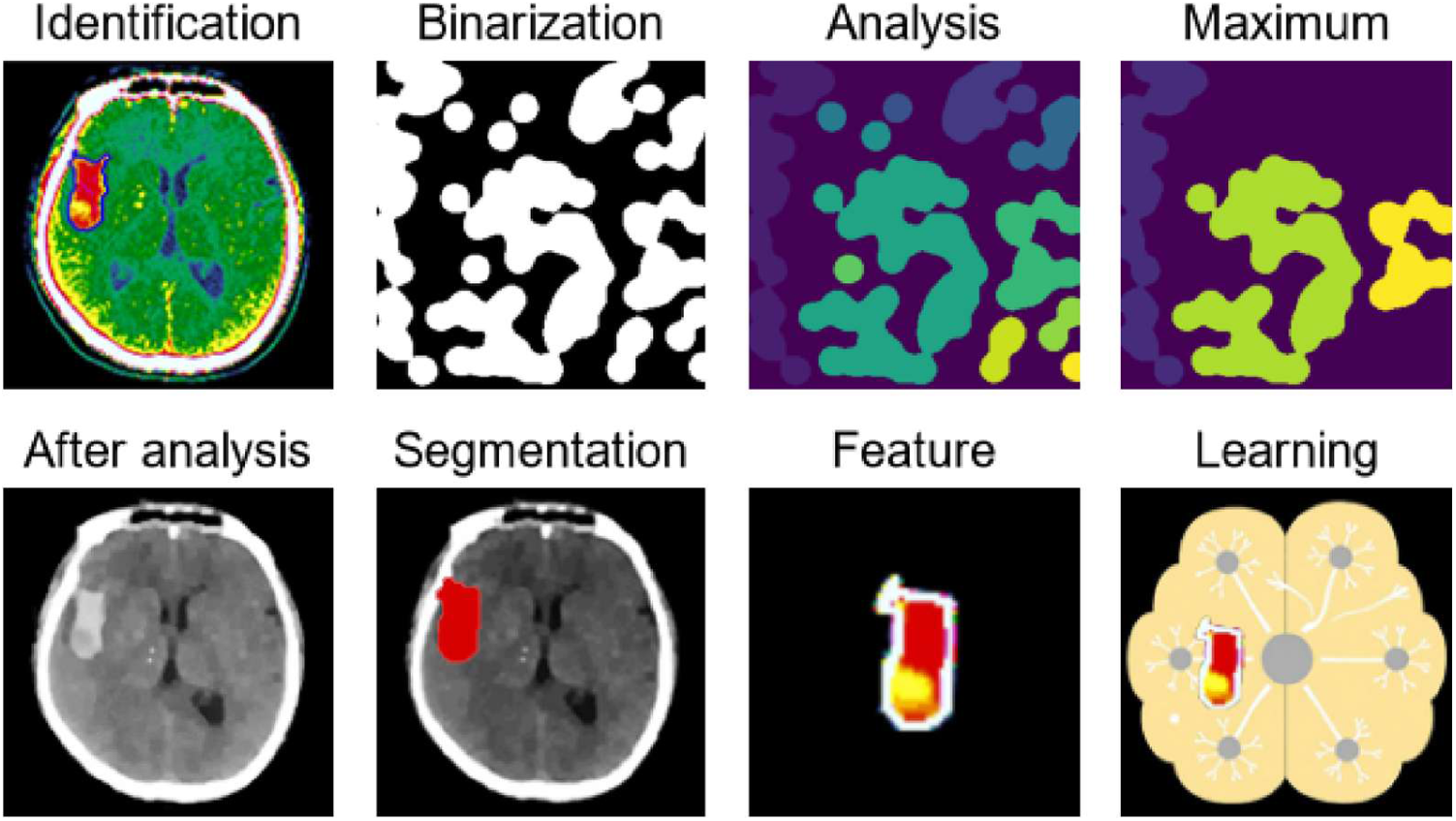
Workflow for pseudocolor-assisted lesion segmentation and radiomic feature extraction. This integrated pipeline outlines the process from original NCCT image to quantitative feature extraction. The sequence includes (A) original NCCT image, (B) pseudocolor enhancement for initial annotation review, (C) manual annotation and quality control by expert neuroradiologists, (D) automated deep learning-based segmentation, (E) generation of a three-dimensional hemorrhage volume of interest (VOI), and (F) extraction of high-dimensional radiomic features, including first-order, shape, and texture features, from the segmented VOI using the PyRadiomics library.

**Sup Fig 2:**
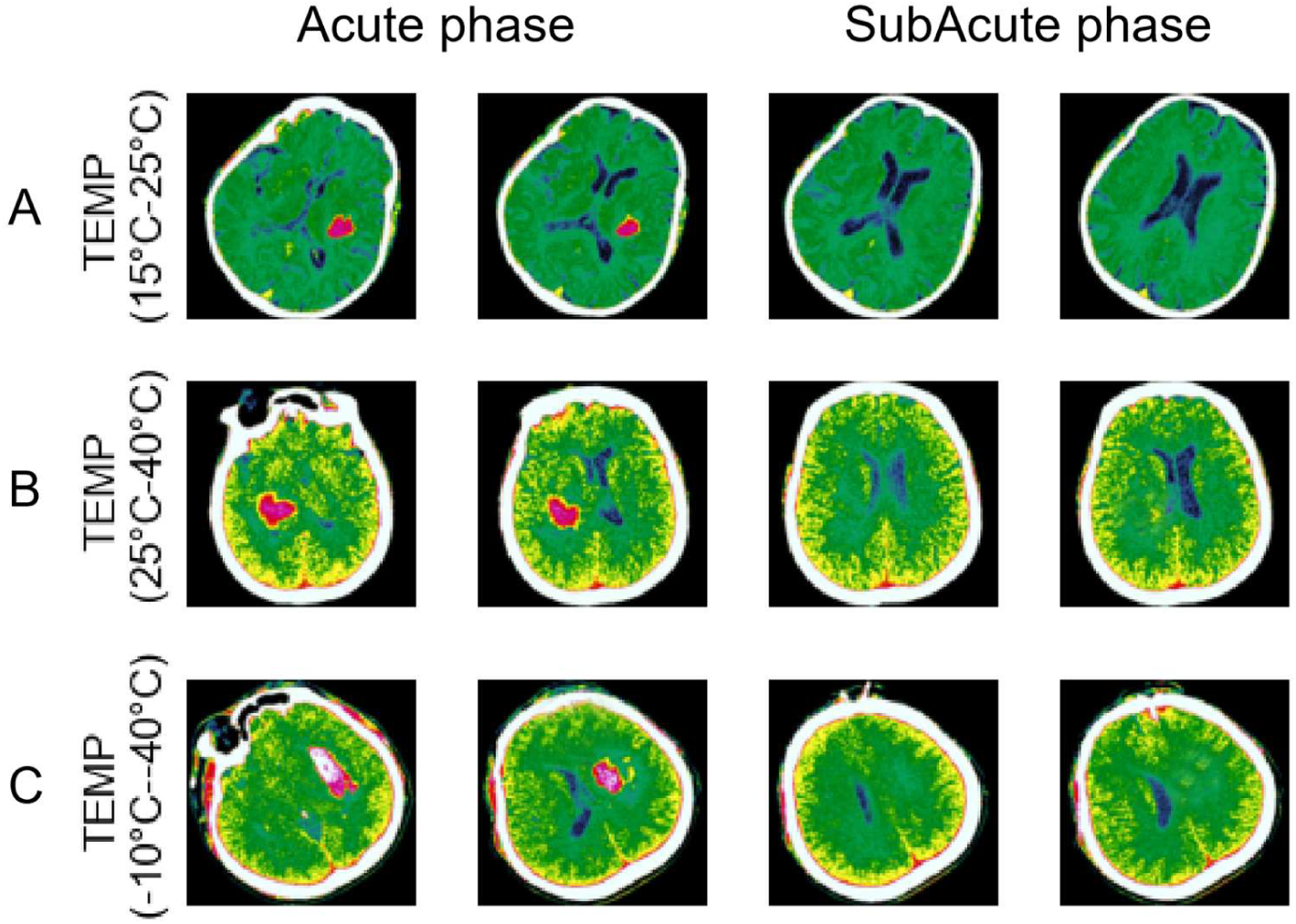
Relationship between ambient temperature at symptom onset and ICH lesion characteristics. The figure compares imaging parameters and clinical outcomes across three predefined ambient temperature groups: Group A (15-25°C), Group B (25-40°C), and Group C (−10 to −40°C). (A) Representative NCCT images at the acute and subacute phases show that Group C exhibited larger lesion volumes and higher imaging intensity, indicating a greater initial injury burden and slower hematoma absorption. (B) Bar charts quantify the mean lesion volume and changes in hematoma absorption rate across the three groups during follow-up, with Group C demonstrating the most severe injury and the slowest recovery trajectory.

**Sup Fig 3:**
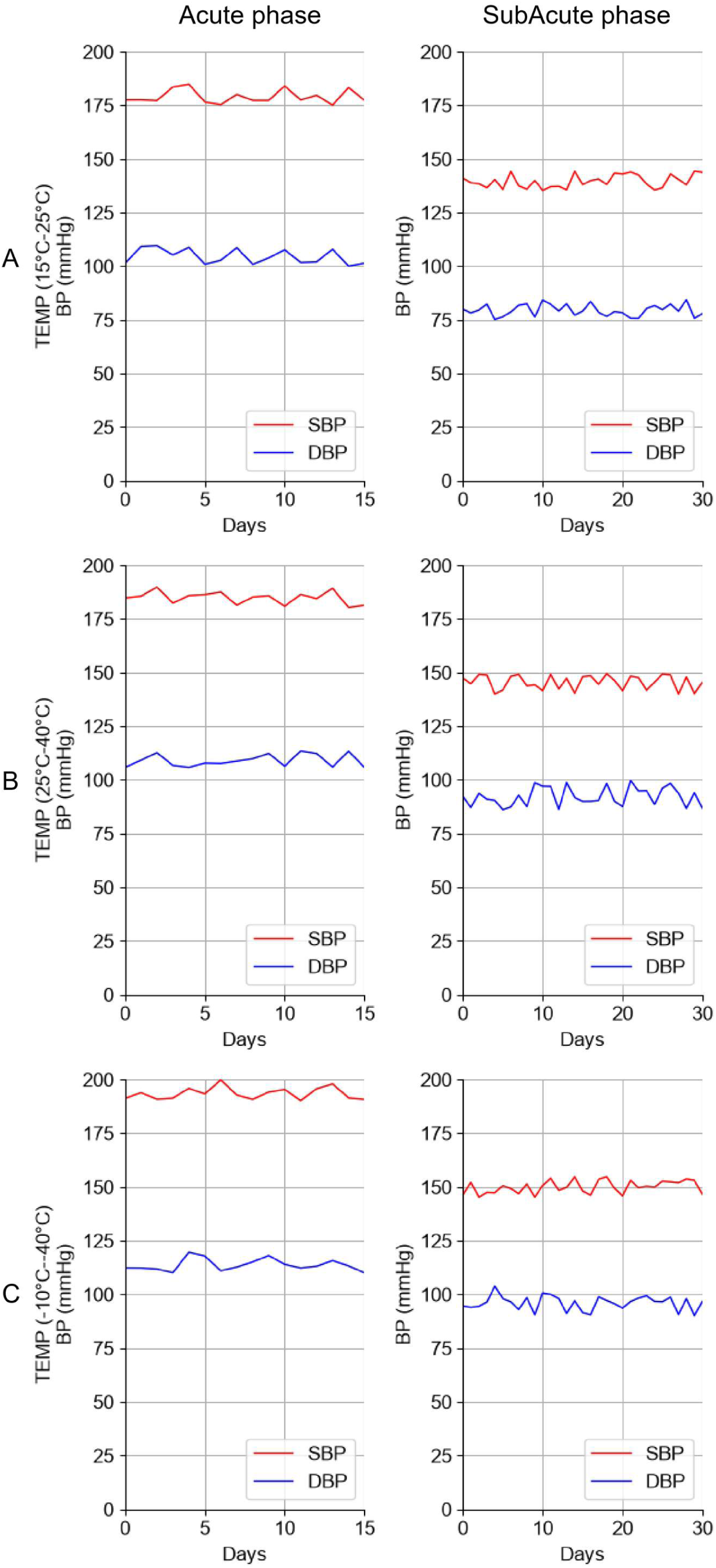
Blood pressure dynamics in relation to ambient temperature groups. The line graphs illustrate the temporal trends in systolic blood pressure (SBP) from admission through the subacute phase for the three temperature groups. Patients in Group C (−10 to −40°C) experienced the most pronounced acute SBP elevation and the slowest SBP recovery during the subacute period. Regression analyses show similar slopes across groups but a clear gradient in SBP values, suggesting that the magnitude of temperature fluctuation may influence blood pressure variability and ICH severity.

**Sup Fig 4:**
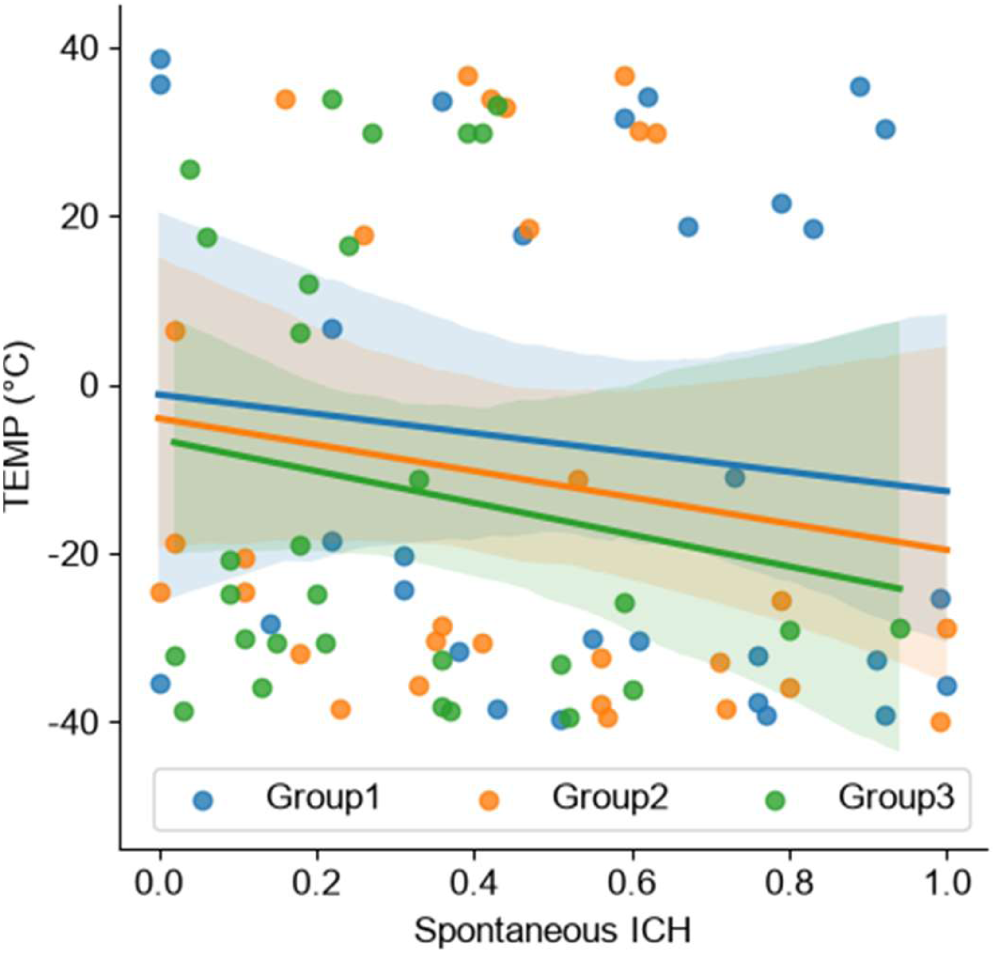
Estimated probability of ICH occurrence as a function of ambient temperature. The bar chart shows the relative risk and estimated probability of ICH occurrence for each temperature group. A clear gradient is observed: P (−10 to −40°C) > P (25-40°C) > P (15-25°C), suggesting that greater temperature fluctuation is associated with a higher likelihood of spontaneous ICH. These findings are hypothesis-generating and support the need for prospective epidemiological studies to validate the potential link between environmental temperature variability and ICH risk.

## References

1. Seiffge, DJ, Fandler-Höfler, S, Du, Y, Goeldlin, MB, Jolink, WMT. (2024) Intracerebral haemorrhage - mechanisms, diagnosis and prospects for treatment and prevention. Nat Rev Neurol, 20, 708–723.10.1038/s41582-024-01035-w

2. Sheth, KN (2022) Spontaneous Intracerebral Hemorrhage. N Engl J Med, 387, 1589–1596.10.1056/NEJMra2201449

3. Puy, L, Parry-Jones, AR, Sandset, EC, Dowlatshahi, D, Ziai, W. (2023) Intracerebral haemorrhage. Nat Rev Dis Primers, 9, 14.10.1038/s41572-023-00424-7

4. Murthy, SB (2024) Emergent Management of Intracerebral Hemorrhage. Continuum (Minneap Minn), 30, 641–661.10.1212/con.0000000000001422

5. Morris, NA, Simard, JM, Chaturvedi, S (2024) Surgical Management for Primary Intracerebral Hemorrhage. Neurology, 103, e209714.10.1212/wnl.0000000000209714

6. Geest, V, Oblak, JP, Popović, K, Nawabi, J, Elsayed, S. (2024) How much of the variance in functional outcome related to intracerebral hemorrhage volume is already apparent in neurological status at admission? J Neurol, 271, 5003–5011.10.1007/s00415-024-12427-9

7. Steiner, T, Purrucker, JC, Aguiar De Sousa, D, Apostolaki-Hansson, T, Beck, J. (2025) European Stroke Organisation (ESO) and European Association of Neurosurgical Societies (EANS) guideline on stroke due to spontaneous intracerebral haemorrhage. Eur Stroke J, 10, 1007–1086.10.1177/23969873251340815

8. Morotti, A, Boulouis, G, Dowlatshahi, D, Li, Q, Barras, CD. (2019) Standards for Detecting, Interpreting, and Reporting Noncontrast Computed Tomographic Markers of Intracerebral Hemorrhage Expansion. Ann Neurol, 86, 480–492.10.1002/ana.25563

9. Sporns, PB, Psychogios, MN, Boulouis, G, Charidimou, A, Li, Q. (2021) Neuroimaging of Acute Intracerebral Hemorrhage. J Clin Med, 10.10.3390/jcm10051086

10. Greenberg, SM, Ziai, WC, Cordonnier, C, Dowlatshahi, D, Francis, B. (2022) 2022 Guideline for the Management of Patients With Spontaneous Intracerebral Hemorrhage: A Guideline From the American Heart Association/American Stroke Association. Stroke, 53, e282–e361.10.1161/str.0000000000000407

11. Gregório, T, Pipa, S, Cavaleiro, P, Atanásio, G, Albuquerque, I. (2019) Assessment and Comparison of the Four Most Extensively Validated Prognostic Scales for Intracerebral Hemorrhage: Systematic Review with Meta-analysis. Neurocrit Care, 30, 449–466.10.1007/s12028-018-0633-6

12. Weissflog, JS, Keller, EJ, Neymeyer, ML, Morotti, A, Dowlatshahi, D. (2026) Systematic review of commercial artificial intelligence tools for the detection and volume quantification in intracerebral hemorrhage. Eur Radiol, 36, 367–395.10.1007/s00330-025-11834-4

13. Hu, P, Yan, T, Xiao, B, Shu, H, Sheng, Y. (2024) Deep learning-assisted detection and segmentation of intracranial hemorrhage in noncontrast computed tomography scans of acute stroke patients: a systematic review and meta-analysis. Int J Surg, 110, 3839–3847.10.1097/js9.0000000000001266

14. Lambin, P, Leijenaar, RTH, Deist, TM, Peerlings, J, De Jong, EEC. (2017) Radiomics: the bridge between medical imaging and personalized medicine. Nat Rev Clin Oncol, 14, 749–762.10.1038/nrclinonc.2017.141

15. Bera, K, Braman, N, Gupta, A, Velcheti, V, Madabhushi, A (2022) Predicting cancer outcomes with radiomics and artificial intelligence in radiology. Nat Rev Clin Oncol, 19, 132–146.10.1038/s41571-021-00560-7

16. Hajiesmailpoor, Z, Kargar, Z, Baradaran, M, Shojaeshafiei, F, Tabnak, P. (2024) Prognostic value of CT scan-based radiomics in intracerebral hemorrhage patients: A systematic review and meta-analysis. Eur J Radiol, 178, 111652.10.1016/j.ejrad.2024.111652

17. Morotti, A, Boulouis, G, Dowlatshahi, D, Li, Q, Shamy, M. (2023) Intracerebral haemorrhage expansion: definitions, predictors, and prevention. Lancet Neurol, 22, 159–171.10.1016/s1474-4422(22)00338-6

18. (Accessed 1 Jun 2019) The International Council for Harmonisation. ICH E8(R1) guideline. 2019.https://www.ich.org/products/guidelines/efficacy/article/efficacy-guidelines.html#8.

19. Collins, GS, Reitsma, JB, Altman, DG, Moons, KG (2015) Transparent reporting of a multivariable prediction model for individual prognosis or diagnosis (TRIPOD): the TRIPOD statement. Bmj, 350, g7594.10.1136/bmj.g7594

20. Cuschieri, S (2019) The STROBE guidelines. Saudi J Anaesth, 13, S31–s34.10.4103/sja.SJA_543_18

21. Chilamkurthy, S, Ghosh, R, Tanamala, S, Biviji, M, Campeau, NG. (2018) Deep learning algorithms for detection of critical findings in head CT scans: a retrospective study. Lancet, 392, 2388–2396.10.1016/s0140-6736(18)31645-3

22. Monteiro, M, Newcombe, VFJ, Mathieu, F, Adatia, K, Kamnitsas, K. (2020) Multiclass semantic segmentation and quantification of traumatic brain injury lesions on head CT using deep learning: an algorithm development and multicentre validation study. Lancet Digit Health, 2, e314–e322.10.1016/s2589-7500(20)30085-6

23. Huang, X, Wang, D, Zhang, Q, Ma, Y, Zhao, H. (2022) Radiomics for prediction of intracerebral hemorrhage outcomes: A retrospective multicenter study. Neuroimage Clin, 36, 103242.10.1016/j.nicl.2022.103242

24. Liu, Y, Zhao, F, Niu, E, Chen, L (2024) Machine learning for predicting hematoma expansion in spontaneous intracerebral hemorrhage: a systematic review and meta-analysis. Neuroradiology, 66, 1603–1616.10.1007/s00234-024-03399-8

25. Yuan, C, Wang, M, Jiang, Y, Li, H, Zhan, C. (2025) Prediction of 180-day post-admission’s GOS in patients with spontaneous intracerebral hemorrhage based on explainable machine learning technology. Medicine (Baltimore), 104, e45402.10.1097/md.0000000000045402

26. Pan, B, Li, F, Liu, C, Li, Z, Sun, C. (2024) Predicting functional outcomes of patients with spontaneous intracerebral hemorrhage based on explainable machine learning models: a multicenter retrospective study. Front Neurol, 15, 149493410.3389/fneur.2024.1494934

27. Pei, L, Fang, T, Xu, L, Ni, C (2024) A Radiomics Model Based on CT Images Combined with Multiple Machine Learning Models to Predict the Prognosis of Spontaneous Intracerebral Hemorrhage. World Neurosurg, 181, e856–e866.10.1016/j.wneu.2023.11.002

28. Yang, K, Fang, J, Zhang, X, Bai, Y, Ji, H (2026) Development and validation of interpretable machine learning models to predict 30-day mortality in patients with intracerebral hemorrhage. Sci Rep.10.1038/s41598-026-61923-7

29. Liang, J, Tan, W, Xie, S, Zheng, L, Li, C. (2025) Prediction of etiology and prognosis based on hematoma location of spontaneous intracerebral hemorrhage: a multicenter diagnostic study. Neuroradiology, 67, 1761–1772.10.1007/s00234-025-03661-7

30. Martinez Oeckel, A, Misselwitz, B, Foerch, C (2025) Incidence But Not Severity of Intracerebral Hemorrhage Shows a Strong Pattern of Seasonal Variation. J Am Heart Assoc, 14, e034077.10.1161/jaha.124.034077

31. Zheng, D, Arima, H, Sato, S, Gasparrini, A, Heeley, E. (2016) Low Ambient Temperature and Intracerebral Hemorrhage: The INTERACT2 Study. PLoS One, 11, e0149040.10.1371/journal.pone.0149040

